# Myelonets define spatiotemporal immunosuppressive programs in ovarian cancer

**DOI:** 10.64898/2026.08.26.26361128

**Authors:** Iga Niemiec, Aleksandra Shabanova, Elias Ruuska, Maija Tissarinen, Zhihan Liang, Gayani Anandagoda, Saundarya Shah, Ziqi Kang, Ada Junquera, Matilda Salko, Ulla-Maija Haltia, Anni Virtanen, Anniina Färkkilä

## Abstract

High-grade serous ovarian carcinoma (HGSC) responds poorly to immune checkpoint blockade, partly due to a macrophage-dominated immunosuppressive microenvironment. We integrated single-cell spatial proteomics and spatial transcriptomics across 50 HGSC tumors and applied SPACEstat to resolve higher-order immune communities and their transcriptional programs. We identified six immune community types, with macrophage-dominated Myelonets representing the predominant spatial pattern of immune organisation. In chemotherapy-exposed tumors, Myelonets showed coordinated lipid metabolism–immunosuppression and inflammation–MHC-II macrophage transcriptional programs, with SPP1, C1Q, VEGF, MMPs, and CCL18 linked to immunosuppressive states and fibroblasts emerging as key mediators of macrophage communication. Chemotherapy contracted large Myelonets while increasing CD8+ T-cell organization into Lymphonets. Persistent macrophage dominance within Myelonets was associated with adverse outcomes among patients who achieved a complete response to treatment. Together, we identify Myelonets as clinically relevant, multicellular immunoregulatory niches sustained by spatiotemporally coordinated macrophage programs and stromal crosstalk.

## Introduction

High-grade serous ovarian carcinoma (HGSC) accounts for approximately 70% of ovarian cancer–related deaths, with over 80% of patients presenting at an advanced stage^1,2^. Standard clinical management relies on primary debulking surgery (PDS) followed by platinum-based adjuvant chemotherapy, or neoadjuvant chemotherapy (NACT) followed by interval debulking surgery (IDS) for unresectable disease^3,4^. Despite the integration of poly ADP ribose polymerase (PARP) inhibitors for tumors harboring homologous recombination deficiency (HRD), recurrence remains common, leading to high mortality^5^. Immune checkpoint inhibitors have yielded disappointing response rates in HGSC^6–8^, largely due to complex, poorly characterized immune evasion mechanisms within the tumor microenvironment (TME).

Although HGSC frequently exhibits prominent infiltrates of CD8+ cytotoxic T lymphocytes (CTLs), a feature typically associated with favorable outcomes^9^, infiltrating lymphocytes are often functionally impaired^10,11^. Overcoming this immunosuppression requires addressing innate immune components, predominantly tumor-associated macrophages (TAMs), which constitute the largest immune fraction in the ovarian TME^12^. Rather than acting as simple M1/M2-polarized entities, TAMs exhibit extensive functional plasticity, frequently establishing an immunosuppressive niche that restrains anti-tumor T cell activity^13,14^. Emerging evidence demonstrates that cell abundance alone is insufficient to predict anti-tumor immunity; instead, the spatial topology and physical proximity of CD8+ T cells to myeloid cells govern therapeutic response and immune escape^15^.

It was recently established that the spatial crosstalk of CD8+ T cells and TAMs is organized into discrete cellular architectures: CD8+ T cells aggregate into coordinated "Lymphonets"^16^, whereas TAMs assemble into dense "Myelonets"^15^. Notably, specific myeloid niches induce localized CD8+ T cell exhaustion upon prolonged contact with antigen presented on major histocompatibility complex II (MHC-II) - a classic M1-polarisation marker^15^. However, the precise transcriptional programs driving these spatial cell-cell interaction patterns remain unmapped. Resolving the molecular mechanisms underlying this spatial heterogeneity is critical for designing therapeutic strategies that reprogram immunosuppressive niches into active anti-tumor hubs.

To bridge this gap, we implemented a multi-omic spatial framework integrating spatial proteomics and transcriptomics across a large cohort of HGSC tissue samples. Leveraging our newly developed computational framework, SPACEstat, we mapped single-cell t-CycIF^17^ topologies into spatial cellular interaction networks. We paired these spatial architectural motifs, discovered at the protein level (Lymphonets and Myelonets), directly with spatially resolved transcriptomic programs. This integrative approach resolves the transcriptional determinants underlying spatial myeloid-lymphoid crosstalk and provides a roadmap for biomarker discovery and targeted immunotherapeutic intervention in HGSC.

## Results

### Integrated spatial proteomics and transcriptomics reveal predominant stromal immune cell infiltration

To study the spatial patterns of immune cell infiltration in high-grade serous carcinoma (HGSC), we collected 50 formalin-fixed, paraffin-embedded (FFPE) tumor specimens from 42 patients who participated in the ONCOSYS-OVA prospective clinical trial (NCT06117384). Samples were collected either during the time of primary debulking cytoreductive surgery (PDS), and/or interval debulking surgery (IDS) conducted after first-line NACT-chemotherapy treatment, constituting a cohort of 16 paired treatment-naive (PDS) and chemo-treated (IDS) samples originating from the same patient (n=8 patients); and 34 IDS samples (n=34 patients) (Materials and Methods 1). Detailed clinical characteristics and survival outcomes are shown in Supplementary Table 1.

Whole slide tumor samples were subjected to multi-omics profiling with high-plex cyclic immunofluorescence (t-CycIF) imaging and GeoMx spatial transcriptomics (ST) (Figure 1A). Figure 1B presents an overview of the collected data and the most important clinical characteristics of the patients.

**Figure 1.**
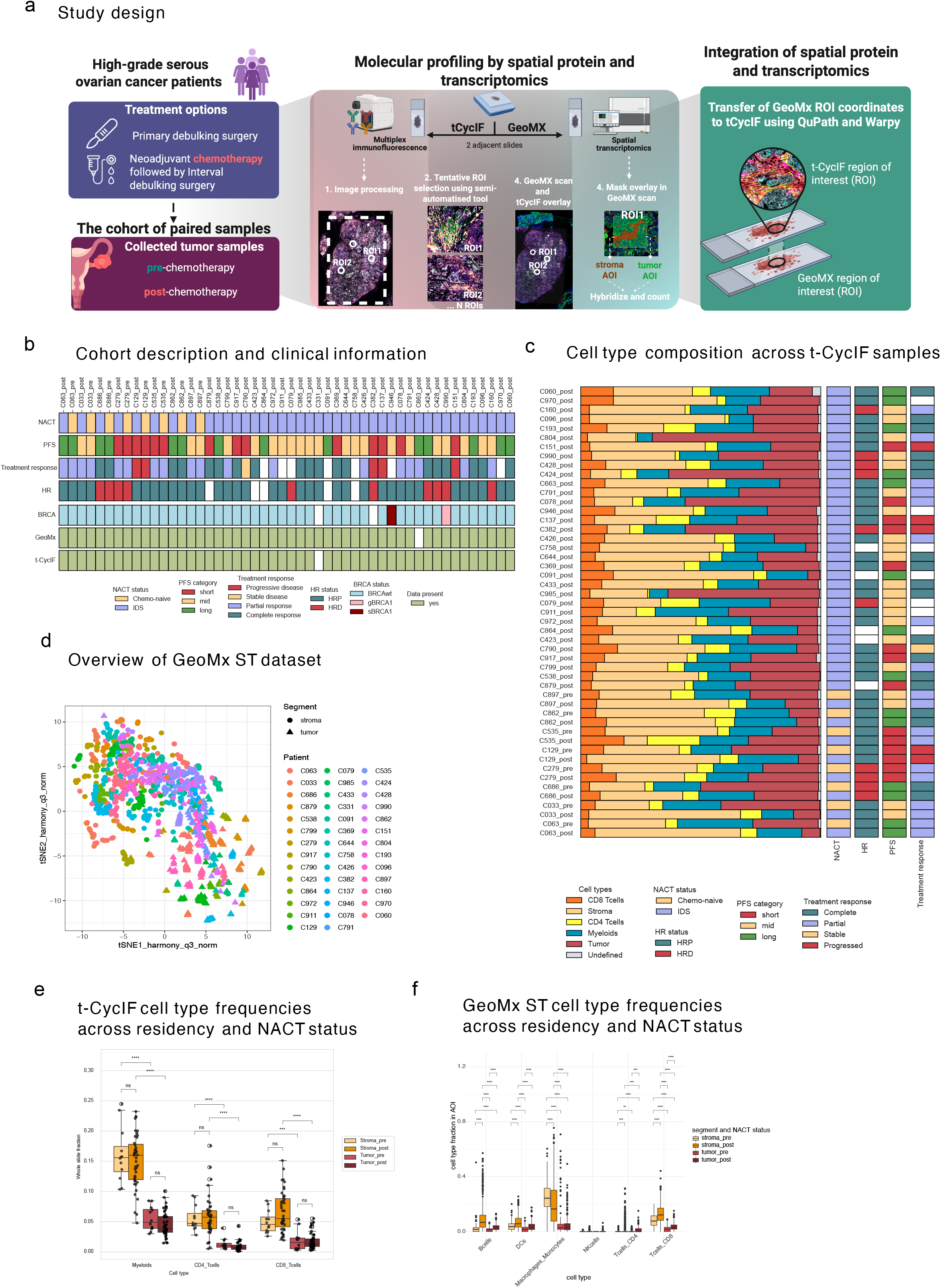
Multi-omics characterization of the high-grade serous ovarian cancer (HGSC) tumor microenvironment (TME) **a.** Schematic overview of samples origin and data and integration workflow. **b.** Data availability and key clinical characteristics of the cohort (n = 50 samples). PFS category was defined based on the lowest and highest 25^th^ percentile within cohort (corresponding to thresholds of 350 days for short PFS group and 602 days for long PFS group). HRD status was defined using ovaHRDscar. **c.** Stacked bar plots of cell type proportions per t-CycIF sample (n = 49 samples) annotated with key clinical characteristics. Cell fractions were calculated as the proportion of each cell type relative to the total number of phenotyped cells in the given sample. **d.** t-SNE projection for AOIs in GeoMx Spatial Transcriptomics data (n=1180 AOIs, n = 49 samples), coloured by patient. Shape corresponds to stroma/tumor AOI segment type. **e** & **f**. Distribution of immune cell fractions between stromal and tumor compartments across samples before (chemo-naive) and after NACT treatment (IDS) in t-CycIF (1E) (n = 12 chemo-naïve (4 duplicates) and n=43 IDS WSIs (2 duplicates)) and GeoMx ST (1F) (n=10 chemo-naïve (2 duplicates) and n=44 samples (3 duplicates)). Cell fractions were calculated as the proportion of each immune cell type relative to the total number of phenotyped cells in the sample (1F) or GeoMx AOI (1G). Asterisks represent significant p-values of paired Wilcoxon rank-sum test between groups (*** - p-value < 0.001, **** - p-value < 0.0001).

On t-CycIF whole-slide image (WSI) data, we phenotyped the following cell types: tumor, stroma, CD8+ T-cells, CD4+ T-cells, myeloid cells, and other cells, and determined their tumor/stromal residency (Figure 1C, Materials and Methods 2.1-2.3). Based on the cells phenotyped in the t-CycIF image, we identified spatial regions of interest (ROIs) containing aggregates of interacting immune cells and found corresponding regions on the adjacent tissue slide (Supplementary Figure 1; Materials and Methods 2.1 and 3.1). Each ROI was further divided into tumor and stromal compartments, which constituted areas of illumination (AOIs), and subjected to GeoMx ST (Materials and Methods 3.2). Employing this protocol, we collected the final ST dataset comprising 1180 AOIs (Materials and Methods 3.3, Supplementary Figure 2). As expected, stromal AOIs from different patients were grouped together in the dimensionality-reduction projection, whereas tumor AOIs clustered separately for each patient, reflecting high inter-patient heterogeneity of HGSC (Figure 1D). Finally, we calculated cell type fractions in each AOI (Materials and Methods 3.5) using a deconvolution algorithm and confirmed that they correspond to the phenotyped cell labels from an analogous region of the adjacent t-CycIF slide (Materials and Methods 3.4, Supplementary Figures 3 and 4).

Similar to our previous findings^15^, the most abundant immune cells were myeloid cells, followed by CD8+ T-cells and CD4+ T-cells. Residency analysis showed that immune cells were more abundant in the stromal than the tumor compartment across both chemo-naive and IDS samples (Figure 1E). Myeloids were the most abundant immune cells in the stroma and had the highest cell fraction in tumor regions. Both CD8+ and CD4+ T cells had similar abundances and were primarily located in the stromal compartments, with substantially smaller fractions located in the tumor regions. Consistent with previous work, we found that overall cell type composition was highly variable across samples, and we did not identify any significant differences in cell type proportions between chemo-naive and IDS samples, nor between samples with long and short platinum-free interval (PFS) groups (Figure 1C). Overall, this analysis showed statistically significant differences in immune cell abundances between the residencies rather than between chemo-naive and chemo-treated samples. ST analysis of the cell-type fraction distributions of stromal and tumor AOIs within the GeoMx ST dataset (Figure 1F) revealed similar proportions and distributions of immune cells. Macrophages, CD8+ T-cells, Dendritic cells (DCs), and B-cells were located preliminarily in stromal compartments, with the most striking difference observed for macrophages.

### Myeloid dominance defines immune cell-type node architecture

To characterize how immune cells are spatially organized within the tumor microenvironment, we applied a spatial analysis framework, SPACEstat, to 55 t-CycIF WSIs from 49 HGSC samples. This cohort comprises 12 treatment-naive samples (8 unique tumors with 4 cross-batch replicate samples from Launonen et al. 2024^15^) and 43 chemo-treated samples (41 unique tumors plus 2 replicate samples from Launonen et al. 2024^15^). For each of the three major immune populations (CD4+ T cells, CD8+ T cells, and Iba1+/CD11c myeloids), spatially clustered cells of the same phenotype were grouped into discrete cell-type nodes (Figure 2A, Materials and Methods 2.4). These cell-type nodes represent previously described Lymphonets^16^ and Myelonets^15^ - interconnected networks of closely interacting lymphoid or myeloid cells. For every cell-type node, the constituent cells were classified as resident in the tumor or the stroma based on tissue-compartment annotations. Each slide was summarized by the fraction of each cell type organized into nodes, the tumor/stroma residency of node-resident cells, the cell-based composition of nodes within WSI, and associations of these features with survival at the patient level.

**Figure 2.**
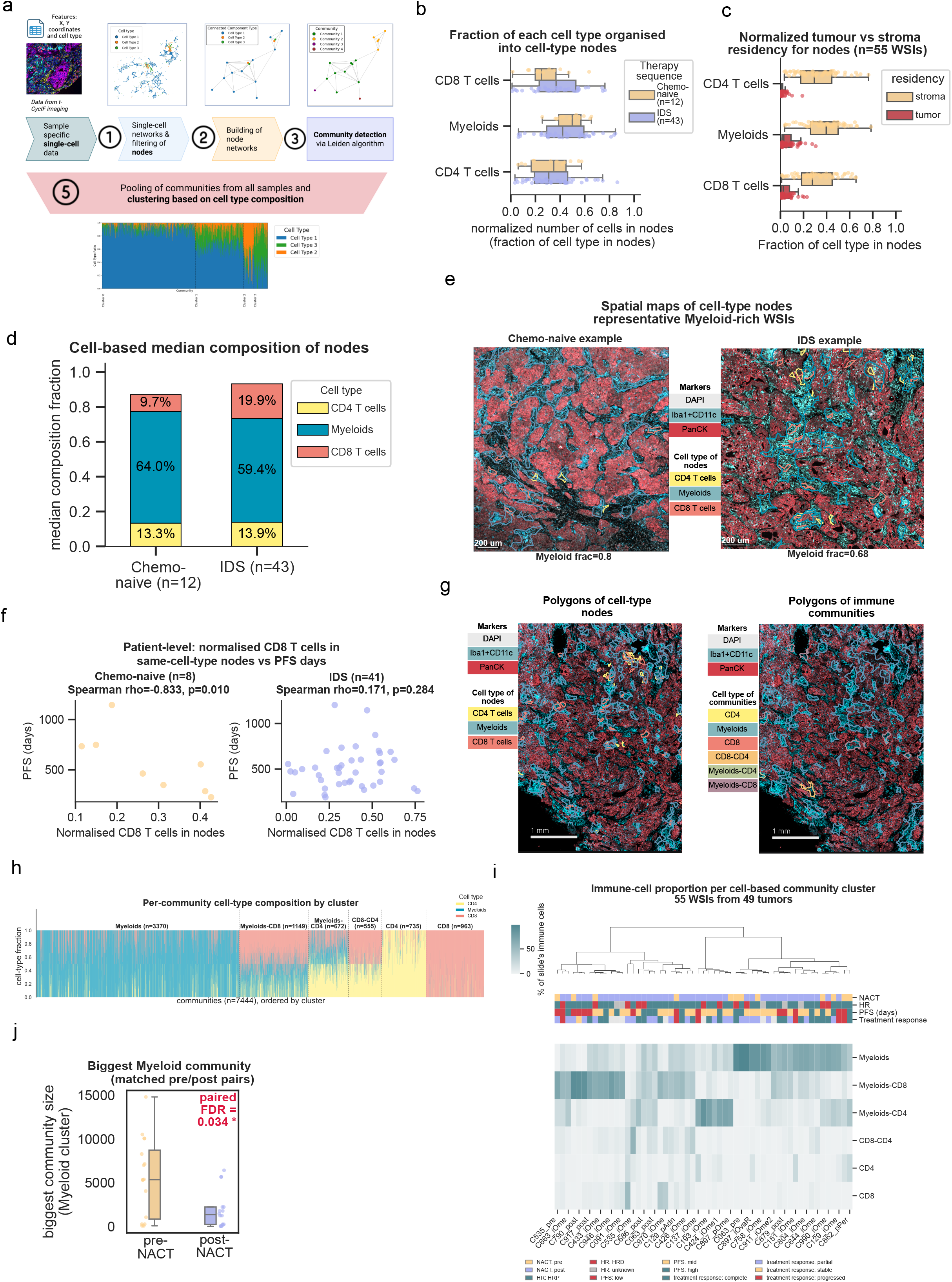
Single cell-type nodes and immune communities **a.** Graph-based workflow for spatial immune community detection and cell composition profiling. (1– 3) Spatial graph pipeline: multiplexed t-CycIF imaging data (cell types and spatial coordinates) is converted into single-cell spatial networks, filtered into nodes of single cell type, and partitioned into discrete spatial immune communities using the Leiden algorithm. (5) Spatial immune communities pooled across samples are clustered by cell type composition. Based on their composition, communities are then grouped into recurring compositional clusters. **b.** Fraction of each immune cell type (CD4+ T cells, myeloids, CD8+ T cells) organized into spatial nodes in chemo-naive (n=12 whole-slide images [WSIs]; 8 unique tumors plus 4 cross-batch replicates) and post-treatment (IDS, n=43 WSIs; 41 unique tumors plus 2 replicates) slides. Boxes mark the median and interquartile range (IQR) with whiskers extending to 1.5× IQR; points represent individual WSIs. Chemo-naive vs. IDS comparisons were performed using a two-sided Mann–Whitney U test with Benjamini– Hochberg FDR correction; no comparison reached FDR < 0.05 (CD8+ FDR = 0.39, myeloids FDR = 0.55, CD4+ FDR = 0.71). **c.** Normalized tumor vs. stroma residency of node-resident cells by cell type: cell counts within tumor- or stroma-resident nodes divided by the total count of that cell type per WSI (n=55 WSIs from 49 unique tumors, therapy sequences pooled). Boxplots as in (b); points represent individual WSIs. **d.** Median cell-based composition of immune nodes in chemo-naive vs IDS samples: for each cell type, the number of cells within nodes of that type is divided by the total number of immune cells across all CD4/ Myeloid/CD8 nodes in the tumor (n=12 chemo-naive and 43 IDS WSIs). **e.** Representative whole-slide spatial maps of single-cell-type nodes in chemo-naive-sample (left) and IDS (right) case. Each panel shows the t-CycIF slide with fluorescent markers DAPI (white, nuclei), Iba1+CD11c (teal, myeloid cells), and PanCK (red, tumour epithelium). Color-coded polygons outline spatially coherent clusters of single cell types (nodes): CD4+ T cells (yellow), Myeloids (green), and CD8+ T cells (salmon), overlaid on the tissue. Both examples were selected for their high myeloid node fraction. Scale bar, 200 μm. **f.** Patient-level association between normalised CD8 T-cell node cell number (CD8 cells in CD8 T-cell nodes / CD8 cells in sample) and progression-free survival in days; one point per patient, replicate tumor samples averaged per patient (chemo-naive n=8, IDS n=41). Spearman correlation per phase: chemo-naive ρ=−0.83, p=0.010; IDS ρ=0.17, p=0.284. **g.** t-CycIF WSI as Figure 2E (DAPI, white; PanCK, red; Iba1+CD11c, teal) with left panel representing single-cell-type nodes, color-coded by cell type and right panel - cell communities, color-coded by cell-based cluster. Scale bar, 1000 μm. **h.** Cell-type composition of individual communities (n = 7,444 communities) grouped by cell-based community clusters. Each column is one community; stacked bars show the fraction of its resident cells belonging to each immune-cell type (CD4 T cells, Myeloids, CD8 T cells; fractions sum to 1). Communities are ordered by the six clusters (Myeloids, Myeloids–CD8, Myeloids–CD4, CD8–CD4, CD4, CD8), with dashed lines marking cluster boundaries and cluster sizes indicated above each segment. **i.** Proportion of each sample’s community-resident immune cells (CD4 T cells, CD8 T cells, Myeloids) assigned to each cell-based community cluster. Rows are the six clusters; columns are individual tumor WSIs (n = 55; 12 pre-treatment, 43 post-treatment). Color scale indicates the percentage of immune cells per WSI. Column color bars annotate therapy sequence (pre/post), HR status (HRP/HRD/unknown), patient PFS split into tertiles (low <25th, mid, high >75th percentile), and treatment response (complete/partial/ stable/progressed). Samples are hierarchically clustered (Ward, Euclidean). **j.** Size of the largest community assigned to the Myeloid cluster in matched pre-/post-treatment sample pairs. Boxplots (median, IQR; whiskers without outliers) with overlaid individual values for n = 16 matched sample-pairs from 8 patients; chemo naive (pre-treatment), IDS = interval debulking surgery (post-treatment). Paired Wilcoxon signed-rank test, FDR = 0.034 (*).

Across all HGSC WSIs, a substantial, cell-type-dependent proportion of each immune population was organized into Lymphonets/Myelonets. On average, myeloid cells showed the highest degree of such organization, with 44.1% (median 44.7%) of cells residing in nodes, compared with 35.3% (median 34.0%) of CD8+ T cells and 33.8% (median 32.1%) of CD4+ T cells; none of these fractions changed significantly between therapy sequences (Figure 2B). Node-resident cells of all three types were mostly stromal in their residency. When normalized to the cells of each type in the whole sample, stromal residency dominated every population (mean 31.8% of CD4+ T cells, 30.6% of CD8+ T cells, and 37.9% of myeloids). In contrast, only 2.0–6.1% of cells of any type resided in tumor-resident nodes (Figure 2C). This indicates that Lymphonets and Myelonets predominantly reside in the stromal compartment, regardless of the sample’s cellular composition. Additionally, we observed that while the sizes of CD8+ and CD4+ Lymphonets were at a median of around 22 and 14 cells, Myelonets can form large structures up to 30–31,198 cells (within a 100um radius used to restrict the maximum size of the immune community). This finding is in line with previous reports describing Lymphonets ranging from 8 to 200 cells and Myelonets ranging from 10 to several thousand cells.

We next examined the cellular composition of immune nodes by calculating the proportion of cells of each cell type among all cells residing in immune nodes. The node composition was dominated by myeloid cells across all tumors: myeloids accounted for 64.0% of node-resident immune cells before treatment and 59.4% after treatment (IDS), whereas CD8+ T cells contributed 9.7% and 19.9%, and CD4+ T cells 13.3% and 13.9% (Figure 2D). Notably, the proportion of CD8+ T-cells roughly doubled in IDS samples (median 9.7% to 19.9%), although not in a statistically significant way (raw p=0.093, FDR=0.158). Myeloids nonetheless remained the single largest cell type node in the majority of samples (12/12 pre-treatment and 37/43 post-treatment; Figure 2E). Because this cell-based metric weights each node by the number of cells it contains, large myeloid aggregates dominate the picture — consistent with myeloids also being the population most frequently organized into nodes (median 44.7%) whereas CD8+ T cells were the least organized before treatment (median 24.9%) and increased their organization into nodes afterwards (median 36.7%; Figure 2B), a shift paralleling the doubling of their node-associated share.

Finally, we examined the association between node-level features and survival outcomes (Supplementary Figure 5). In the pre-treatment group at the patient level, a higher fraction of CD8+ T cells organized into Lymphonets correlated with poorer survival: greater node organization predicted shorter PFS (Figure 2F, ρ = -0.83 for PFS, p = 0.010, n = 8). However, these associations were not significant after false discovery rate correction, and no node features were linked to survival in the post-treatment cohort (n=41).

### Higher-order spatial network analysis uncovers temporal dynamics of multicellular immune communities

To examine higher-level spatial organization, we defined multicellular immune communities as spatially adjacent cell-type nodes within 100 µm (Figure 2A, Materials and Methods 2.5), encompassing both cell-cell interactions and paracrine cytokine communication^18^. It yielded 7,444 immune communities across all tissue sections (Figure 2G). Immune communities were classified into six clusters based on their dominant cell-type composition (Myeloids, Myeloids–CD8, Myeloids–CD4, CD8–CD4, CD4, CD8; Figure 2H). Cluster membership was well balanced at the community level (Myeloids n = 3,370; Myeloids–CD8 n = 1,149; CD8 n = 963; CD4 n = 735; Myeloids–CD4 n = 672; CD8–CD4 n = 555), with the single-cell-type clusters together accounting for the majority of communities.

Median cell-type node-ratio profiling showed that each immune community cluster corresponded to a distinct compositional phenotype (Supplementary Figure 6). The pure Myeloid cluster was the most abundant (3,370/7,444 communities; 45%) and largely defined by single-cell-type in composition (median Myeloids node ratio = 1.00), similar to the other single-immune-type clusters (CD4: 735 communities; CD8: 963), whereas the mixed clusters were more heterogeneous in represented cell types, as expected. Single-immune-type clusters were sharply defined, with a median ratio of 1.00 for their related cell type Lymphonets.

We next assigned each community-resident immune single cell to one of the six cell-based clusters and, for each tumor, computed the percentage of the WSI community-resident immune cells (CD4/CD8/Myeloids, n = 4.76 M of 29.99 M scanned immune single cells) that fell into each cluster. At the tumor level, the proportion of each community type varied markedly across tumors, and clustering resolved the sections into a small number of compositional archetypes dominated by Myeloids (representing large Myelonet communities), Myeloids–CD8-, or Myeloids–CD4-enriched communities, indicating that Myelonets dominate immune community architecture.

Among all immune community features, only the size of the largest Myelonet community changed significantly between matched pre- and post-treatment sections (paired Wilcoxon signed rank; FDR = 0.034; Figure 2J). Across 16 matched treatment-naive – post treatment pairs from 8 patients, the largest Myeloid community contracted approximately fourfold, from a median of 5,350 cells (IQR 846–8,730) before treatment to 1,348 cells (IQR 221–2,220) after treatment (from mean 5,604 to 1,899), indicating a marked shrinkage of the dominant Myelonet community following neoadjuvant therapy.

Similar distributions of immune cell interaction patterns were found among selected ROIs in the ST dataset (n=736 ROIs), clustered based on their immune composition, and averaged for AOIs within each ROI (Materials and Methods 3.6). This analysis revealed five different types of ROIs (Figure 3A), with the most abundant type composed almost exclusively of Macrophages and representing Myelonet communities (n=268), followed by a cluster of ROIs dominated by Macrophages and CD8+ T-cells (n=252). Remaining ROI types displayed mixed communities with overrepresentation of other immune cells (Th17, Tregs, NKs) (n = 96), CD4+ T-cells (n = 74), and B-cells (n = 46). We confirmed that ROI clusters correctly represent immune communities identified in t-CycIF analysis by demonstrating very high concordance between these two types of labels for analogous regions of adjacent slides (Materials and Methods 3.4, Supplementary Figure 7). Distribution of ROI clusters between chemo-naive and IDS samples revealed the same pattern as observed for t-CycIF communities, with Macrophage-dominated ROIs (Myelonet communities) and Macrophage-CD8 ROIs being a dominant immune community in chemo-naive and IDS samples, respectively (Supplementary Figure 8).

**Figure 3.**
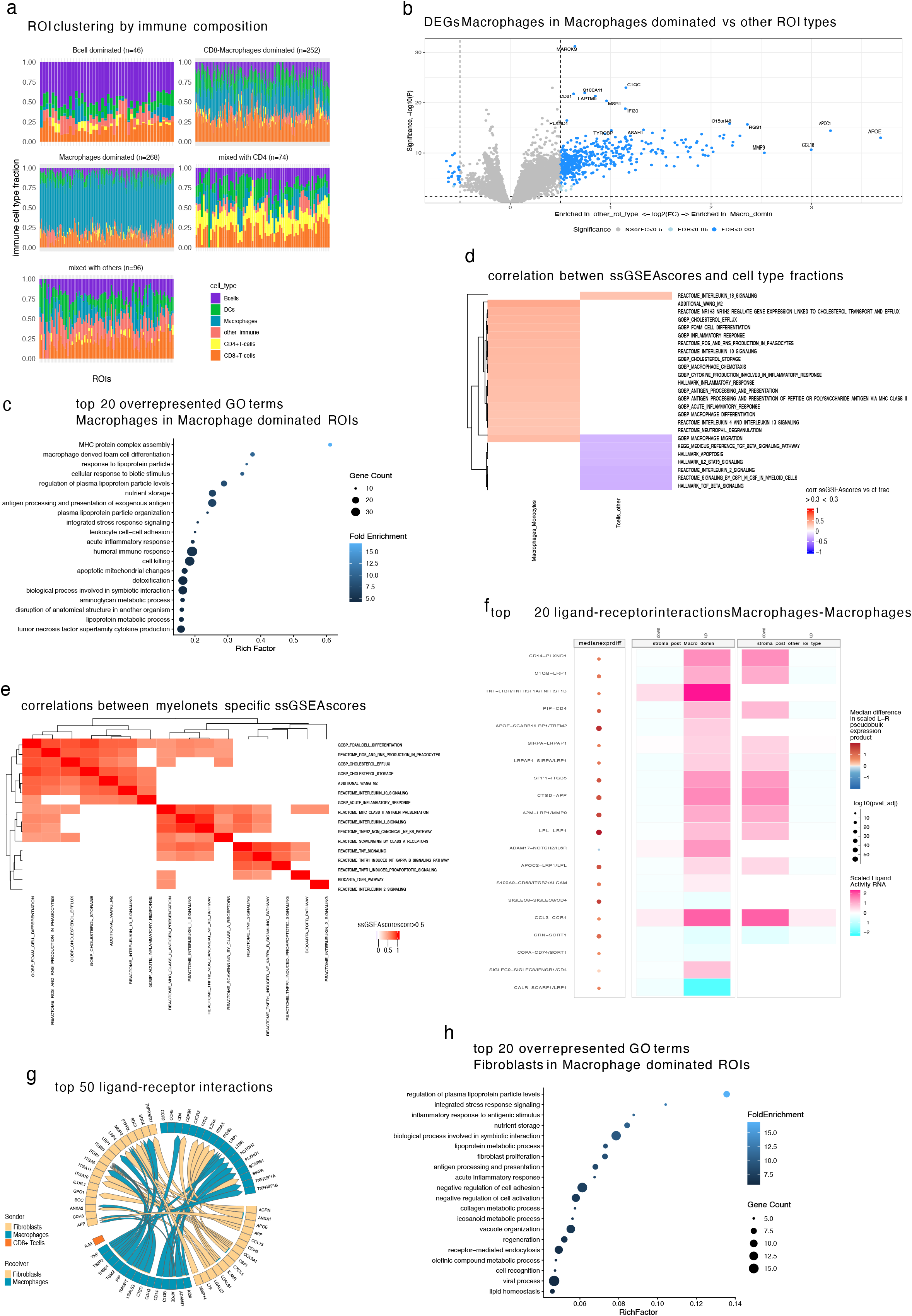
Transcriptomics characterization of Myeloid-dominated immune communities **a.** Stacked barplots with immune cell composition across clusters of GeoMx ST ROIs (n=736 ROIs, n=49 samples). Fractions of immune cells obtained with SpatialDecon deconvolution algorithm were summed for AOIs within ROI. Relative immune fraction was calculated as fraction of given immune cell divided by total fraction of all immune cells within given ROI. Clustering was performed with GMM method with n=5 clusters based on relative immune fractions of selected cells (Bcells, DCs, Macrophages, CD4+ T-cells, Cd8+ T-cells and other immune cells). The number of ROIs belonging to each cluster is indicated in the bracket. **b.** Volcano plot showing Differentially Expressed Genes between stromal AOIs belonging to Macrophage dominated ROI cluster (n = 177 AOIs) vs stromal AOIs of all other clusters (n = 278 AOIs) within each IDS sample containing at least one AOI belonging to Macrophage dominated ROI (n=34 samples). DEGs were calculated using LMM model with SampleID as cofounder, based on Macrophage specific deconvoluted transcriptomic profiles obtained with BayesPrism deconvolution algorithm. Dark blue color represents significantly over/underexpressed genes (FDR < 0.01, log2FC > abs(0.5)). **c.** Bubble plot with top 20 overrepresented GO terms calculated based on significantly overexpressed DEGs (n=485 genes, FDR <= 0.01, log2FC >= 0.5) from stromal AOIs of IDS samples belonging to Macrophages dominated ROI cluster (n=177 AOIs, n=34 samples), calculated for Macrophage specific deconvoluted transcriptional profile. Significant GO terms (n = 980 terms) were clustered by the similarity to their parent term (thr = 0.5), filtered to remove all the parent terms, ranked by Rich Factor and filtered to one highest ranked GO term per similarity cluster to represent only unique terms. **d.** Heatmap showing Spearman correlation coefficients between ssGSEA score for selected immune-related pathways (n=256 pathways) and AOI cell type fraction within all stromal AOIs of IDS samples (n=575 AOIs, n=44 samples). SsGSEA scores were calculated based on Macrophage specific deconvoluted transcriptional profiles. Only pathways and cell types displaying the Spearman correlation coefficient higher than 0.3 or lower than –0.3 are displayed. **e.** Heatmap showing Spearman correlation coefficients between ssGSEA scores for selected pathways representing key transcriptional features of Macrophage-dominated immune communities (n=18 pathways) within stromal AOIs of IDS samples belonging to Macrophages dominated ROI cluster (n=177 AOIs, n=34 samples). SsGSEA scores were calculated based on Macrophage specific deconvoluted transcriptional profiles. Only values of Spearman correlation coefficient higher than 0.5 are displayed. **f.** Top 20 ligand-receptor interactions between Macrophages within stromal AOIs of IDS samples belonging to Macrophages dominated ROI cluster (n=177 AOIs, n=34 samples) vs stromal AOIs of all other clusters from the same samples (n = 398 AOIs, n=34 samples). Interactions were calculated with MultiNichenetR algorithm based on Macrophage specific deconvoluted transcriptional profiles grouped by ligand and ranked by prioritization score. Heatmap represents scaled ligand activity scores for each ligand-receptor pair and bubble plot represents median difference in scaled L-R pseudobulk expression product, with positive values indicating overexpression in AOIs belonging to Macrophages dominated cluster. **g.** Chord plot representing top-50 prioritized ligand receptor interactions between Macrophages, CD8+ T-cells and Fibroblasts within stromal AOIs of IDS samples belonging to Macrophages dominated ROI cluster (n=177 AOIs, n=34 samples). Empty lines represent ligands/receptors prioritized in stromal AOIs of IDS samples belonging to all other clusters. Interactions were calculated with MultiNichenetR algorithm based on deconvoluted transcriptomic profile of each analyzed cell type. **h.** Bubble plot with top 20 overrepresented GO terms calculated based on significantly overexpressed DEGs (n=105 genes, FDR <= 0.01, log2FC >= 0.5) from stromal AOIs of IDS samples belonging to Macrophages dominated ROI cluster (n=177 AOIs, n=34 samples), calculated for Fibroblast specific deconvoluted transcriptional profile. Significant GO terms (n = 552 terms) were clustered by the similarity to their parent term (thr = 0.5), filtered to remove all the parent terms, ranked by Rich Factor and filtered to one highest ranked GO term per similarity cluster to represent only unique terms.

### Myelonets host coordinated immunosuppressive and inflammatory macrophage states

Since Myelonet communities constitute the most common immune colocalization pattern, we aimed to further analyze whether the transcriptomic profile of Macrophages in Myelonet communities differs from that of Macrophages in contact with other immune cells (Materials and Methods 3.8, Supplementary Table 3). We found that in stromal regions of IDS samples, macrophages in Myelonet communities were overexpressing genes connected with monocyte recruitment, positive regulation of chemotaxis, migration, and cell-cell adhesion signatures (chemokines: CCL5, CCL2, CXCL4/CCL20, CXCR4), suggesting active formation of Myeloid communities (Figures 3B and 3C).

The highest overexpressed genes within Myelonet communities were Apolipoprotein E (APOE) and Apolipoprotein C-I (APOC1) (log2FC > 3) (Figure 3B), being part of overrepresented signatures connected to lipid and cholesterol transport and storage, steroid metabolism, response to lipoprotein particle (along with Lipoprotein lipase - LPL), foam cells differentiation, and stress response (Figure 3C). Additionally, we found overexpression of component 1q (C1Q) proteins (C1QA/C1QB/C1QC) as part of signatures indicating negative regulation of immune response and connected with fatty-acid metabolism reprogramming^19^. That suggests immunosuppressive behavior in Myelonet communities, as APOE+ and C1Q+ Macrophages have been linked to the promotion of an immunosuppressive environment across different cancer types, including ovarian cancer. We also found overrepresentation of pathways associated with the acute inflammatory response (especially the interleukin-1 [IL-1] and tumor necrosis factor alpha [TNFα] cytokines), along with antigen processing and presentation on MHC-II (Figures 3B and 3C), suggesting Macrophage activation.

At the same time, we identified several genes and signatures indicating immunosuppressive properties of Macrophages within Myelonet communities: M2 polarization (CXCL8/CCL3/CCL5)^22,23^, pro-cancer behavior (Secreted Phosphoprotein 1 - SPP1, CCL20 chemokine)^24,25^, response to Transforming Growth Factor beta (TGFβ) and IL-10, angiogenesis (Vascular endothelial growth factor - VEGF) and Extracellular Matrix (ECM) degradation (metalloproteinases: MMP9, MMP19)^26,27^. One of the most overexpressed genes (log2FC = 3) was the immunosuppressive CCL18 chemokine, a marker of M2 TAMs, linked to helper T-cell type 2 (Th2) switching and to the promotion of angiogenesis^19^ (Figures 3B and 3C).

We next analyzed correlations between the activity of selected immune pathways (Supplementary Table 4) in Macrophages and the corresponding AOI cell-type fractions (Materials and Methods 3.7). We identified a positive correlation between the fraction of Macrophages and single-sample gene set enrichment analysis (ssGSEA) scores for inflammatory response, M2, antigen processing and presentation (APP) on MHC-II, chemotaxis, activation, differentiation, IL-10, IL-4/IL-13 signaling (promoting M2 polarisation^29^), and VEGF signatures in all stromal AOIs of IDS samples. At the same time, we found negative correlations between IL-2 and TGFβ signaling and fractions of other T-cells (regulatory T-cells and Th17). Frequencies of other cell types were not found to influence the activity of immune signatures of Macrophages. Thus, we confirmed that disrupted lipid metabolism, immunosuppression, and inflammatory response are directly influenced by an elevated number of Macrophages in spatial proximity, rather than being an artifact of ROI clustering.

### Myelonet macrophages couple lipid dysregulation with immunosuppression and inflammation

Next, we aimed to further characterize relationships between the main identified transcriptional features of Myelonet communities within stromal compartments of IDS samples: APOE and lipid metabolism, SPP1-integrin communication, inflammatory response to IL-1, IL-2, and TNFα, response to immunosuppressive TGFβ and IL-10, M2-polarisation, APP on MHC-II, and VEGF signaling (Supplementary Table 4). First, we confirmed that 18 selected pathways representing these processes sufficiently characterize Myelonet communities by performing hierarchical clustering of all stromal AOIs from IDS samples based on their activity and observing a separate cluster of Myelonet communities (Supplementary Figure 9). Next, we analyzed correlations between selected pathway activity scores, calculated from Macrophage-specific deconvoluted transcriptional profiles. We identified two main transcriptional programs, consisting of highly correlated pathways (Spearman correlation coefficients of 0.5-0.9): lipid metabolism-immunosuppression (lipid/stress/IL-10/M2) and inflammation-MHC-II (TNFα/IL-1/MHC-II).

The first identified intercorrelated transcriptional program consisted of signatures related to foam cell differentiation and cholesterol efflux and storage (driven by APOE/APOC1), reactive oxygen/nitrogen species (ROS/RNS) production, IL-10 signaling, M2-polarisation, and acute inflammatory response. This interesting finding suggests that dysregulated lipid metabolism relates to cellular stress and immunosuppressive Macrophage behavior. Another cluster of interconnected signatures was linked to IL-1 and TNFα signaling (especially through Tumor necrosis factor receptor 2 - TNFR2), scavenger receptor activity (related to C1QA/B/C), and APP on MHC-II, suggesting Macrophage activation in response to inflammatory molecules. Interestingly, some of these processes were also highly correlated with foam cell formation and ROS production, providing a link between these two main transcriptional programs.

This analysis suggests that the transcriptional features specific for Macrophages within Myelonet communities are functionally connected and form two separate transcriptional programs of lipid metabolism-immunosuppression (lipid/stress/IL-10/M2) and inflammation-MHC-II (TNFα/IL-1/MHC-II). This finding confirms that the observed aberrant spatial plasticity of Macrophages is a wide-ranging process profoundly changing their metabolism and overall phenotype.

### Macrophage–fibroblast crosstalk supports immunosuppressive features of Myelonet communities

Our analysis identified several important transcriptional features of stromal Myelonet communities in IDS samples, concentrated around two main programs: lipid metabolism and inflammation. To determine whether cellular communication affects the observed traits, we used a modified MultiNicheNetR framework (Materials and Methods 3.9) to infer Ligand–Receptor communication among three main cell types within these spatial communities: Macrophages, fibroblasts, and CD8+ T-cells (which constitute 5-15% of immune cells within Myelonet communities).

Analysis of top-prioritized ligand–receptor pairs within Macrophage–Macrophage communication (Supplementary Table 5, Figure 3F) revealed that Macrophages’ cellular crosstalk directly drives several previously identified processes. We identified several important ligand–receptor pairs associated with lipid metabolism and lipoprotein-related signaling, including APOE–SCARB1/LRP1/TREM2, APOC2–LRP1/LPL, C1QB–LRP1, and A2M–LRP1/MMP9. The TNFα transcriptional program was also reflected in macrophage communication, as we identified strong signaling through TNF–TNFRSF1A (TNFR1)/TNFRSF1B (TNFR2) (Figure 3F). That suggests an active role for Macrophages as both producers of this highly inflammatory protein and recipients of this signaling, resulting in the regulation of distinct downstream transcriptional programs, as revealed by correlation analyses. In addition, we identified several other ligand–receptor pairs connected to immunosuppressive Macrophages populations: SPP1–TGB5^20^, SIGLEC9– SIGLEC8/IFNGR1/CD4^21^, and SIRPA–LRPAP1, suggesting impaired phagocytic activity^22^. Finally, the main chemokine signaling between Macrophages was found to be driven by CCL3-CCR1 (Figure 3F). All the identified pairs are highly specific to Melonets. They are characterized by high downstream activity, strengthening the hypothesis that lipid- and TNFα-related communication is directly connected to Macrophages and is a key feature of this spatial niche, alongside certain markers of immunosuppressive Macrophage populations (C1Q, SPP1, SIGLEC9) (Figure 3F).

Interestingly, CD8+ T-cells were not found to be active participants in multicellular crosstalk within myeloid-dominated spatial niches, as analysis of the top 50 prioritized ligand receptor interactions did not identify any ligand–receptor pairs associated with this cell type (Supplementary Table 5, Figure 3G). On the contrary, communication between fibroblasts and macrophages is very active within Myelonet communities (Figure 3G). We found that lipoprotein-related signaling is also highly active in reciprocal macrophage– fibroblast communication, as we identified the APOE–LRP1/SCARB1 interaction among the top-prioritized pairs. Interestingly, the most active pair in fibroblast–macrophage communication was colony-stimulating factor 1 (CSF1) − SIRPA/CSF3R, suggesting that fibroblasts might be an important source of this potent chemoattractant and M2-switch facilitator^22,23^.

Our finding on the role of fibroblasts in immunosuppressive activity of macrophages was further confirmed by our additional differentially expressed genes (DEGs) and Gene Ontology (GO) enrichment analysis of fibroblasts in Myelonet communities (Figure 3H). We found that Fibroblasts residing in these spatial niches were overexpressing genes linked to angiogenesis, lipid transport, storage, and catabolism (FABP3/APOE/APOC1), response to wounding, TGFβ, and ROS. In contrast, fibroblasts in other ROI types overexpressed multiple pro-inflammatory factors (including CCL13, CXCL5, CXCL9, CCL19, and IL1B). Overall, our findings suggest that fibroblasts in Myelonet communities are under cellular stress, exhibit highly active lipid metabolism, and are actively engaged in immunosuppressive communication with macrophages.

### Post-chemotherapy Macrophage-rich Myelonets associate with poor clinical outcomes

To test whether the Myeloid-program activity observed in the tumor microenvironment is reflected across the whole-slide t-CycIF architecture rather than confined to individual ROIs, we related identified Myeloid-specific transcriptional programs (lipid metabolism-immunosuppression and inflammation-MHC-II pathways’ ssGSEA scores) measured on the matched stroma AOI to Myeloid aggregation on the adjacent t-CycIF section (Figure 4A, Materials and Methods 3.10, Supplementary Table 4). On the 652 matched node– AOI pairs (n=49 from 46 unique tumor samples (7 chemo-naïve & 39 chemo-treated tumors), a random-intercept-per-sample linear mixed model showed that the summed cell count of Myeloid-labeled components within the ROI, adjusted for other-immune aggregation, was positively associated with two functionally defined Myeloid-specific transcriptional programs (FDR q<0.05 for all readouts) (Figure 4B). The lipid metabolism-immunosuppression program (lipid-il10-m2, lipid-il10-m2_dge) increased by +0.034–0.036 SD per IQR increase in aggregated Myeloid cells (95% CIs 0.018–0.052; q<5×10⁻⁵), and the inflammation-MHC-II program (IL1-TNFR2-M2) by +0.018–0.035 SD per IQR across its readouts (TNFα the weakest, q=0.018). In contrast, the remaining readout (IL2-VEGF) was unrelated to Myeloid aggregation (q=0.76) (Figure 4B, Supplementary table 6). Activity of both Myeloid-specific programs therefore rises proportionally to the size of Myeloid communities within the matched ROIs across the aligned WSI, indicating a program-specific link between Myeloid communities and their transcriptional readout in the tumor microenvironment rather than a generic Myeloid-burden effect.

**Figure 4.**
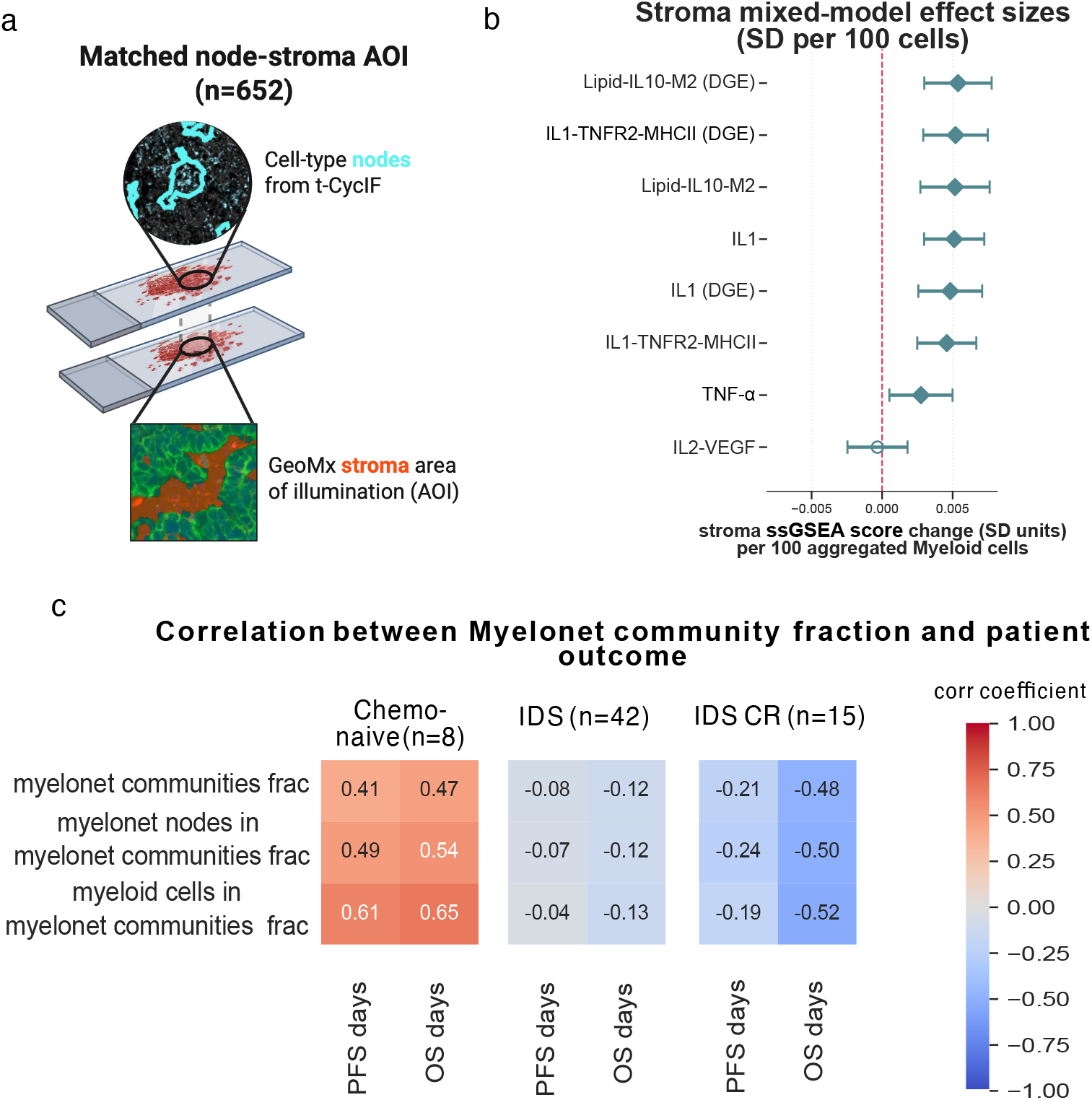
Whole slide spatial mapping of Myelonet-specific transcriptional programmes and their association with survival outcome **a.** t-CycIF on an adjacent section resolved single cells into cell-type nodes which were overlaid onto GeoMx ROIs by whole-slide affine registration (Methods and Materials 3.4), and specifically to stroma areas of illumination which supplied Myeloid-specific transcriptional programmes’ ssGSEA scores. **b.** Linear mixed model with a random intercept per sample (z-score ∼ (sum of Myeloid component sizes)/100 + (sum of other-immune component sizes)/100) fitted on the 652 stroma-assigned node-AOIs (n= 49 from 46 unique tumor samples) for each Myelonet-specific transcriptional programmes’ ssGSEA scores. Each pathway score was z-standardized (mean 0, SD 1) across the scored stroma AOIs, so coefficients are representing the expected change in standard-deviation (SD) units per 100 aggregated Myeloid cells, adjusted for other-immune aggregation. Points and whiskers show the model coefficient and its 95% CI (coefficient ± 1.96 SE); filled diamonds mark pathways surviving Benjamini–Hochberg FDR (q<0.05), open circles mark non-significant pathways; the dashed line marks the null (zero effect). Pathways ordered by descending coefficients. **c.** Spearman correlation coefficient between OS/PFS in days and: 1) number of Myeloid communities divided by total number of communities per sample, 2) number of myeloid cell-type nodes cells belonging to Myelonet community cluster divided by total number of myeloid cell-type nodes per sample, 3) number of myeloid cells belonging to Myelonet community cluster divided by total number of myeloid cells per sample. Calculated separately for each group of samples: Chemo-naive (n=8), IDS (n=42), IDS originating from patients with complete response (CR) to the primary treatment (n=15).

Finally, we investigated whether the proportion of macrophages within Myelonet communities is directly associated with patients’ clinical outcomes. We calculated the proportion of myeloid cells/myeloid cell-type nodes that belong to Myelonet communities, relative to the total number of myeloid cells/myeloid cell-type nodes. We found very small negative correlations between progression-free survival/overall survival (PFS/OS) and Myelonet cells/nodes/clusters for all analyzed IDS samples (n=42 samples) (Figure 4C). Interestingly, among patients with a complete response to first-line treatment (n=15), we found a significant negative correlation between OS and Macrophage abundance in Myelonet communities (Spearman correlation coefficients: –0.48, –0.5, and –0.52 for communities, nodes, and cells, respectively). Interestingly, the same correlation calculated for treatment-naive samples (n=8) revealed a strong positive correlation between Myelonets and patient survival (Spearman correlation coefficient > 0.6). This suggests that the negative impact of Myelonets depends on the intrinsic features of the tumor, possibly linked to dying tumor cells following a complete response to chemotherapy.

## Discussion

Our integrative spatial multi-omics analysis enabled us to study both the overall spatial patterns of immune interactions and the underlying transcriptional programs in HGSC. Utilizing a network-based spatial analysis approach, we identified previously undescribed higher-level multicellular communities. We uncovered Myelonet communities as a main pattern of spatial immune localization within HGSC TME. We demonstrated that Myelonet communities of chemo-treated samples are functionally distinct niches characterized by two interconnected transcriptional macrophage programs of lipid metabolism-immunosuppression and inflammation-MHC-II, revealing coordinated macrophage plasticity within the tumor microenvironment. We identified SPP1, C1Q, VEGF, MMPs, and CCL18 connected with elevated immunosuppressive properties of Myelonet communities and a crucial role of fibroblasts in macrophage intercellular communication and regulation. Finally, we showed an association between Myelonet communities and negative clinical outcomes in HGSC patients with a complete response to primary treatment.

Consistent with the previous reports, our systematic network approach quantified that both lymphoid and myeloid cells form Lymphonets and Myelonets - interconnected groups of spatially interacting cells^15,16^, localized predominantly in stroma. Clustering of immune communities by their cell type composition revealed six different types of spatial immune interaction patterns with Myeloid (representing large Myelonets) and Myeloid-CD8 being the most abundant, constituting almost 45% and 15% of all immune communities, respectively. While the distribution of immune communities between chemo-naive and IDS samples was similar, we found that Myelonet communities’ size is significantly larger in chemo-naive samples and regressed after chemotherapy treatment. Interestingly, the number of CD8+ T cells forming Lymphonets increased after chemotherapy. Together, these findings indicate that chemotherapy reshapes not only immune-cell abundance but also their spatial connectivity, shifting the stromal immune architecture from large, interconnected myeloid niches toward smaller myeloid communities accompanied by increased spatial co-organization of CD8+ T cells.

We found that the transcriptomic profiles of macrophages in Myelonet communities are distinct from those in other communities, but only within the stromal compartments of IDS samples, confirming our previous observations that chemotherapy profoundly affects this immune community. We described three main transcriptional features of macrophages within Myelonet communities: significant alterations in lipid metabolism, strong inflammatory response mediated by TNFα, and M2-like immunosuppressive properties. We demonstrated that these features are spatially coregulated by the number of macrophages, form two distinct interconnected transcriptional programs (lipid metabolism-immunosuppression and inflammation-MHC-II), and rely on macrophage and fibroblast cellular communication.

Altered lipid metabolism and communication, driven by high overexpression of APOE and APOC1, were primarily connected to lipoprotein communication, lipid and cholesterol transport, and foam cell differentiation. APOE+ TAMs were linked to promoting an immunosuppressive environment, T-cell exhaustion, anti-PD1 resistance, and poor prognosis in numerous cancer types^20–23^. Multiple other studies link disruptions in TAM’s lipid metabolism to poor prognosis. For example, OC cells were found to promote excessive cholesterol efflux in TAMs directly, leading to overexpression of fatty acid-binding proteins (FABP) 4 and 5 (also overexpressed in Myeloid-dominated communities) and to poor prognosis. Ligand receptor pairs connected to APOE signaling in macrophages-macrophages and macrophages-fibroblast cellular communication, especially involved low density lipoprotein receptor-related protein 1 (LRP1) and triggering receptor expressed on myeloid cells 2 (TREM2) receptor on Macrophages-TREM2 is recently gaining a lot of attention as a marker of poor prognosis in multiple cancer types and its role in multiple processes such as phagocytosis, lipid metabolism, cell survival, and inflammatory responses is currently being investigated^20^. Importantly, a recent study by Ghisoni et al. 2025^26^ conducted on mouse models of recurrent OC identified wide networks of TREM2/APOE-high and TAMs as a main immunosuppressive component of the TME of BRCA wild-type tumors. Our finding that the activity of lipid-related, ROS/RNS production, acute inflammatory response, M2 and IL-10 signaling signatures form a distinct interconnected transcriptomic profile, suggests that altered lipid metabolism, cellular stress, and immunosuppressive behavior might arise from the same underlying processes, likely connected to acute inflammatory response.

Another important Myelonet-specific transcriptional program was the inflammatory response mediated by TNFα and IL-1, directly connected to macrophage communication and mediated by the TNF-TNFRSF1A/B ligand-receptor pair. TNFα and IL-1 signaling signatures formed a distinct transcriptional program along with antigen processing and presentation on MHC-II. Both features are usually linked to M1-like polarisation^27^, although no production of other classical M1 cytokines (IL-1b, IL-12, IL-6, CCL8, CXCL9, CCL10)^20^ apart from TNFα was observed, suggesting that a proinflammatory environment might result rather in elevated cellular stress (indicated by apoptotic and stress response signatures) than a typical M1 response. In line with this hypothesis, we identified several additional features of Myelonet communities that are linked to immunosuppressive TAM populations within HGSC. We identified high overexpression of genes linked to M2 polarization (including highly overexpressed CCL18)^19^, pro-cancer behavior (SPP1, CCL20)^22,23^, angiogenesis (VEGF), ECM degradation (MMP9, MMP19)^24,25^, and negative regulation of immune response (C1QA/B/C). Many of which were identified as crucial in macrophage-macrophage communication within Myeloid-dominated communities (eg A2M–MMP9, C1QB–LRP1, SPP1–ITGB5, SIGLEC9–SIGLEC8). SPP1/CXCR9 ratio was found to be a better indicator of pro-tumoral TAMs polarization in HGSC than traditional M2 markers^23^. In the same study, SPP1 correlated with expression of CXCL5, CXCL8, IL1A, IL1B, IL1RN, TNFα signaling in TAMs and CD8+ T-cells, and TGFβ signaling in monocytes and CD8+ T-cells^21^, all of them (apart from IL1A) being overexpressed in Macrophages in Myelonet communities. In addition, multiple studies found C1Q+ SIGLEC9+ and MMP9+ TAMs populations being associated with immunosuppression, immune escape and chemotherapy resistance in ovarian cancer^22–25^. Another study identified immunosuppressive TAMs populations, characterized by joint TREM2, C1Q, SPP1 and APOE overexpression^26^ suggesting the existence of one underlying mechanism of immunosuppression. The hypothesis of immunosuppressive phenotype of Myeloid communities is also in line with the previous findings of M2-polarised macrophages being linked with spatial CD8+ T-cell exclusion^15,27^. We propose that macrophages in Myelonet communities display a high level of cellular stress and altered lipid metabolism which might drive their pro-cancer behavior regardless of the strong inflammatory response and traditional M1/M2 polarization patterns.

Interestingly, the connection between Myeloid communities and patient outcome differs profoundly between chemo-naive and IDS samples and among IDS samples with different responses to chemotherapy. While a higher amount of Myeloid cluster is correlated with better survival in chemo-naive samples, this effect is reversed in IDS samples, especially these with complete response to chemotherapy. This might suggest that the observed immunosuppressive features of macrophages within Myelonet cluster are triggered by the presence of large amounts of dying tumor cells and subsequent inflammation, possibly accounting for the relapse of the patients. External dataset validation and functional modeling are needed to assess and uncover potential causal relationships between these processes, as well as between Myeloid communities and clinical outcomes in HGSC patients.

## Materials and Methods

### Resource availability

#### Lead contact

Further information and requests for resources and reagents should be directed to and will be fulfilled by the lead contact, Anniina Färkkilä.

### Experimental model and study participant details

#### Cohort description

High-grade serous carcinoma (HGSC) tumor samples and clinical data were collected as part of the ONCOSYS-OVA NCT06117384 clinical trial, approved by the Ethics Committee of the Helsinki University Hospital (HUS334/2021). For triaging the patients to undergo neoadjuvant chemotherapy (NACT), ESMO-ESGO 2019 guidelines^4^ were utilized. In accordance with the ethical standards of the 1975 Declaration of Helsinki, every patient in the ONCOSYS-Ova trial provided informed written consent to the collection, storage, and analysis of samples and subsequent data.

This study utilizes a total number of 50 tumor specimens, collected from HGSC patients (n=42) at the time of laparoscopy or primary debulking surgery (PDS) (chemo-naive) and interval debulking surgery (IDS). Sample characteristics and patients’ clinical data are summarized in Supplementary Table 1.

Samples were grouped into short-medium-long progression-free survival (PFS) and overall survival (OS) groups based on thresholds corresponding to 0.25 and 0.75 quartiles of the overall distribution of these values across the whole cohort. The threshold values were 350 and 602 days for short and long PFS, respectively, and 613 and 1083 days for short and long OS, respectively.

### Method details

#### 1. Study design

A total of 50 formalin-fixed, paraffin-embedded (FFPE) HGSC tumor specimens (from 42 patients) were analyzed in this study. The cohort comprises 16 paired samples – collected for the same patient during both chemo-naive and chemo-treated (IDS) (n=8 patients); and 34 IDS samples (n=34 patients).

From each sample, 2 adjacent slides of 5um thickness were cut. One of them was subjected to tissue high-plex immunofluorescent (t-CycIF) imaging, while the other was subjected to GeoMx spatial transcriptomics (ST) analysis. A subset of the cohort comprising n=16 GeoMx slides (10 paired pre- and post-NACT samples and 6 IDS samples) has been previously analyzed and published in Launonen et al. 202^15^. For these samples, we obtained a freshly cut slide, adjacent to the GeoMx slide from the other side of the sample, and subjected it to t-CycIF staining. These paired adjacent tissue slides were used for t-CycIF-guided GeoMx region of interest (ROI) selection (see Materials and Methods 3.1) and analyzed jointly.

#### 2. Highplex t-CycIF imaging

##### 2.1 Highplex t-CycIF imaging and image processing

All FFPE tissue sections were scanned with a RareCyte CyteFinder scanner following the t-CycIF protocol, following one of five validated antibody panels: a 13-plex panel A (n=16 samples from Launonen et al. 2024^15^), a 13-plex panel B (n=14), an 18-plex panel C (n=5, including 4 duplicate samples from Launonen et al. 2024^15^ tumor blocks), a 20-plex panel D (n=2, including 1 duplicate sample from Launonen et al. 2024^15^ tumor blocks), or a 39-plex panel E (n=21, including 3 duplicate samples). Samples with their corresponding antibody panels are listed in Supplementary Tables 1 and 2.

Raw image data were processed using a custom pipeline (https://github.com/farkkilab/image_processing). Briefly, image tiles were stitched using ASHLAR^28^. Whole-slide images were subsequently segmented with Mesmer (DeepCell framework, v0.12.10)^29^ based on the first DAPI channel from the first t-CycIF cycle. Protein marker intensities, both raw and white top-hat-filtered, were quantified as the mean intensity per nuclear mask. Low-quality cells and artifacts were filtered out using CyLinter^30^ (executed through all steps before log-transformation). Finally, signal intensities were normalized across batches using UniFORM^31^, with a manually selected representative sample for each protein marker. Throughout the process, Napari^32^ was used for visual assessment.

Following image processing and quality filtering, three slides were excluded due to technical issues: C129_post (panel A), C331_post (panel B), and C686_post (panel D). In total, 55 t-CycIF WSIs representing 49 unique tumor samples (along with 6 cross-batch technical duplicates) were utilized for downstream analysis. This cohort comprises 12 treatment-naive WSIs (8 unique tumors plus 4 cross-batch replicate samples from Launonen et al., 202415) and 43 post-chemotherapy/IDS WSIs (41 unique tumors plus 2 replicate samples). Replicate slides were included across analytical runs to verify cross-batch consistency and are displayed as individual image-level data points across relevant figures.

##### 2.2 t-CycIF cell type phenotyping

Single-cell phenotyping was performed using a multi-step consensus workflow combining rule-based clustering, threshold-based gating, and refinement rules. Initial cellular annotations were assigned using Tribus^33^ applied to arcsinh-transformed (1/5) normalized intensity values using a hierarchical logic table (Supplementary Table 7) with specific parameters (depth=3, σ=1, learning_rate=1, clustering_threshold=100, undefined_threshold=0.0005, and other_threshold=0.4). Marker intensity features were selected based on pre-processing: PanCK, Iba1, and stroma markers (α-SMA or Vimentin, depending on panel availability) were measured using raw intensities, whereas CD11c, CD4, and CD8a were measured using white top-hat-filtered intensities to suppress background noise. To refine single-cell identities, channel-specific manual expression gates were set using scimap (scimap.pl.gate_finder), guided by initial automated estimates (Otsu thresholding) and interactive visual inspection on image overlays. Finally, initial Tribus annotations were refined using a strict three-rule logic to prevent misclassification: (1) tumor cells (Tumor_Tumor) positive for the CD8a gate were re-annotated as cytotoxic T cells (Tumor_Immune_CD8_Tcells); (2) immune-annotated stromal cells (Stroma_Immune_X) lacking expression of the corresponding immune marker (X) were demoted to Stroma_Stroma only if positive for structural markers (α-SMA or Vimentin); and (3) immune-annotated tumor cells (Tumor_Immune_X) lacking expression of marker X were demoted to Tumor_Tumor only if positive for PanCK. Ambiguous cells lacking confirmation of both immune and structural/epithelial markers retained their initial annotation. Across all samples, six primary cell lineages were quantified: tumor cells, stromal cells, CD4+ T cells, CD8+ T cells, CD11c+ and IBA1+ myeloid cells.

##### 2.3 t-CycIF spatial analysis

###### 2.3.1 Tissue images and null models

For the single-cell-level analysis of the t-CycIF data, a binary raster image of the tissue was created for each sample at an initial resolution of 1 µm/pixel. Each pixel was initialized as False and set to True if it corresponded to the centroid of a segmented cell. This point mask was then dilated by 5 iterations using scipy.ndimage.binary_dilation^34^, and the resulting image was downsampled by aggregating pixels into 10 µm × 10 µm bins, with a bin set to True if any of its constituent 1-µm pixels were True.

Null models representing complete spatial randomness (CSR) were generated for each sample by using the pixels of the tissue raster as the pool to sample coordinates for a homogeneous Poisson point process. Pooling from the downsampled pixels assured spacing between the generated cells. For each cell type, the observed density (cells per pixel) was estimated by dividing the observed cell count by the tissue-mask area. A simulated cell count for that type was then drawn from a Poisson distribution with mean equal to this observed density multiplied by the tissue area so that, in expectation, the simulated count matches the observed count, with added Poisson variability. Coordinates for the simulated cells were sampled without replacement from the tissue-mask pixel pool, and cell labels were randomly assigned based on the individual counts of each cell type drawn from the Poisson distribution.

###### 2.3.2 Tumor-stroma residency

Tumor residency was determined by first estimating a smoothed density map of tumor cells for each sample. Tumor-cell centroids were binned into a 20 µm × 20 µm grid restricted to the defined tissue region, and the resulting count grid was smoothed with a Gaussian kernel using scipy.ndimage.gaussian_filter^34^ with sigma parameter set to 2, producing a continuous kernel density estimate of tumor-cell density. The same rasterization and smoothing procedure was applied to 30 independent CSR-simulated samples generated with matched cell-type densities for each corresponding tissue sample (section 2.3.1).

An empirical, one-sided Monte Carlo p-value was then computed for each pixel. Pixels with a p-value of 0.05 or lower were classified as tumor regions. Connected components smaller than 10 pixels were removed, and the resulting binary tumor region was further restricted to pixels within the tissue image. Each cells’ tumor residency was then assigned by binning its coordinates to the same grid resolution and checking whether the coordinates fell within the binary tumor region.

###### 2.3.3 Tumor-stroma interface

For the interface analysis, tumor and stroma regions were defined independently using the same rasterization and Gaussian-smoothing procedure described in Section 2.3.2 (cell centroids binned to a grid restricted to the tissue footprint, followed by Gaussian smoothing). Unlike the tumor-residency regions, the tumor and stroma regions here were defined by direct thresholding of the smoothed density map with a threshold of 0.1. Connected components smaller than 10 pixels were excluded from both binary regions.

Each region was then dilated by 1 iteration to ensure edge overlap, and a boundary band ("rim") was extracted from each by eroding the region using scipy.ndimage.binary_erosion^34^ with 4 iterations and subtracting it from the dilated region. The tumor-stroma interface was defined as the intersection of the tumor and stroma rims. Cells were classified as interface cells if their binned coordinates fell within this intersection. The distance from each cell to the interface was then computed using its original unbinned coordinates. The distance was defined as the Euclidean distance to the nearest interface-classified cell, found using scipy.spatial.cKDTree^34^.

###### 2.3.4 Gini coefficient of cell type density and nearest neighbor distances

Nearest neighbor distances between cells that were used to calculate the mean nearest neighbor distances between cell types were calculated using scipy.spatial.cKDTree^34^.The Gini coefficient of cell density of the cell types was calculated by dividing the sample into a regular 100 x 100 grid and calculating the number of cells in each grid cell. Empty cells were excluded. The Gini coefficient was calculated from the distribution of cell counts across occupied grid cells.

##### 2.4 t-CycIF Defining Lympho- and Myelonets

###### 2.4.1 Single cell-type nodes

To define Lympho- and Myelonets, cell-type-specific single-cell networks were constructed. Sample-specific networks were constructed based on the x and y coordinates of single cells. In the constructed networks, single cells acted as nodes, and edges were defined between nodes based on a distance threshold (section 2.4.2). Cells that had Euclidean distance smaller than the distance threshold were connected with an edge. From the resulting single-cell networks, connected nodes were extracted. All network operations were done using the Python package NetworkX^35^.

Myelonets were defined as connected nodes of myeloid cell single-cell networks with an equal or larger number of nodes than a defined myelonet-specific cell-counts threshold (section 2.4.2). The two types of Lymphonets (CD8 and CD4) were similarly defined as connected parts of CD8 T cell and CD4 T cell single-cell networks that had a larger number of nodes than cell count thresholds separately tuned for CD8 and CD4 T cells.

###### 2.4.2 Tuning single cell-type nodes’ parameters

The cell type-specific distance thresholds and cell count thresholds for the analysis were chosen by running the defining of Myelo- and Lymphonets with all combinations of distance thresholds in the range of 3 - 30 µm and cell count thresholds in the range of 2 - 40 for each analyzed cell type in each observed sample and ten null model samples simulated from that sample. For each sample, the optimal distance and cell count threshold combination was determined as the one maximizing S, where:

*S=(x-y)e^{-py},*

x is the number of cells in Myelo- and Lymphonets identified in the observed sample, y is the mean number of Myelo- and Lymphonets identified across the ten simulated null model samples, and p is a penalty parameter controlling the influence of the expected number of cells in Myelo- and Lymphonets under the null model. This metric favors parameter combinations that maximize the number of observed Myelo- and Lymphonets while penalizing combinations that lead to a high number of Myelo- and Lymphonets in the simulated null model samples. The value of p was set to 5.0 × 10⁻⁵ as this value was observed to result in a good balance between prioritizing the number of cells in Myelo- and Lymphonets in observed samples and penalizing it in simulated null models. The final threshold parameter combinations for each cell type were acquired by taking the floored average of both values from optimal pairs across all samples. The final distance thresholds were 24 µm for myeloid cells, 28 µm for CD8 T cells, and 29 µm for CD4 T cells. The final cell count thresholds were 30 for myeloid cells, 13 for CD8 T cells, and 9 for CD4 T cells.

##### 2.5 Defining immune communities

Lympho- and Myelonets were used to construct sample-specific higher-order networks, with edges drawn at a distance threshold of 100 microns, chosen to match cytokine diffusion distancesof cytokines^18^. An edge was drawn between myelonet-myelonet, lymphonet-myelonet, or lymphonet-lymphonet pair if between the two networks there existed a cell pair with a smaller Euclidean distance than the distance threshold. Network construction was performed using the Python package NetworkX^35^.

The Leiden algorithm^36^, implemented in the Python package leidenalg, was used to partition the higher-order network into immune communities. The algorithm was run with the constant pots model (CPM) as the objective function and the distance between nodes as edge weights. The algorithm was run with the resolution parameter of 1.0 × 10⁻⁵ and 100 iterations. Communities comprising fewer than 20 cells were excluded.

Communities partitioned from the connected single-cell-type networks were clustered into six clusters based on their cell-type composition. Clustering was performed using k-means clustering from the Python package scikit-learn^37^.

The tumor-stroma residency of Lympho- and Myelonets was determined by the majority residency of the single cells comprising them. Similarly, the tumor-stroma residency of the immune communities was determined by the majority residency of the Lympho- and Myelonets comprising the communities

#### 3. GeoMx DSP Spatial Transcriptomics

The GeoMx dataset utilized in this study consists of 320 areas of illumination (AOIs) from 16 samples, published in Launonen et al. 202^15^, and an additional 1088 AOIs originating from 42 samples collected and processed as part of this study. The GeoMx DSP spatial transcriptomics procedure was conducted according to the prescribed GeoMx guidelines for slide staining and scanning. Tissue slides chosen to perform GeoMx sequencing were adjacent to the one on which t-CycIF was performed, to enable guided ROI selection. Data was processed using a custom pipeline (https://github.com/farkkilab/geomx-processing).

##### 3.1 GeoMx ROI selection with t-CycIF crop overlay

t-CycIF stainings (Materials and Methods 2.1) were employed to guide the selection of regions of interest (ROIs, n=713) in the GeoMx experiment. The chosen regions contained aggregates of immune cells stained for CD8 (CD8+ T cells), CD4 (CD4+ T cells), IBA1, and CD11c (myeloid cells), to identify ROIs with different immune cell-type colocalization patterns. The adjacent slide subjected to the GeoMx procedure was stained for PanCK, Vimentin, and CD45 to visualize tumor, stroma, and immune cells, respectively. The list of all antibodies used can be found in Supplementary Table 2.

ROIs were pre-selected on adjacent t-CycIF tissue sections utilizing a semi-automated selection process. ROIs from adjacent t-CycIF images were selected based on four immune markers: CD4, CD8a, CD11c, and Iba1. The raw lower-resolution images (pyramid level 3; ZARR 3, 2.6 µm/pixel) were pre-processed. First, a binary matrix for each marker was created by thresholding channel intensity at the 85th–90th percentile, adjusted per sample. Next, morphological operations were used to minimise small noise and merge signal into larger clusters: (1) opening with a 10 × 10 pixel (26 × 26 µm) kernel; (2) closing with a 20 × 20 pixel (52 × 52 µm) kernel; (3) dilation with a 20 × 20 pixel (52 × 52 µm) kernel. Overlaps of the pre-processed images were then found for every combination of two, three, and four markers. Each overlap map was post-processed to identify larger connected neighborhoods (8-connectivity component labeling), expanded to merge nearby regions (distance of 400 pixels, 1.04 mm), and filtered to keep only the largest regions (those above the 90th percentile of region size); the ten largest ROIs per combination were saved. These ROIs were then checked for spatially overlapping duplicates across combinations, assigning a label to each ROI with the largest number of markers, and annotated for PanCK positivity. For samples with limited tissue area or fold artifacts (n=3), selection was repeated on the usable tissue region. Finally, all candidate ROIs were visually inspected and selected in Napari^32^. In total, we obtained ten ROIs per sample in batch 2 and a sample-dependent number in batch 3; in each sample, we attempted to collect as many diverse ROIs as possible. The final ROIs were saved as rectangles overlaid on composite PNG images generated from the t-CycIF WSI (DAPI, PanCK, and Vimentin channels) after 90° rotation.

The chosen t-CycIF image with candidate ROIs was imported into the GeoMx software and manually superimposed onto the GeoMx scan using landmarks present in both images to identify regions corresponding to pre-selected ROIs. Consistency in choosing similar types of areas, avoiding tissue folds, necrotic regions, or adipose tissue, while maintaining similarities in ROI size and cell count across all samples, was aimed at. ROIs were selected within the tumor, stromal, or tumor-stromal interface (TSI). Additional separation for tumor and stromal compartments (constituting different AOIs) has been performed using masks generated manually within GeoMx DSP software.

##### 3.2 GeoMx slides handling and library preparation

The GeoMx spatial transcriptomics procedure was conducted in the Färkkilä laboratory. All samples were handled according to the prescribed GeoMx guidelines for slide staining (GeoMx DSP Manual Slide Prep User Manual, MAN-10150-06), scanning (GeoMx DSP Instrument User Manual, MAN-10152-06), and library preparation (GeoMx DSP NGS Readout User Manual, MAN-10153-06).

The slides were baked at 60°C for one hour and sequentially deparaffinized and rehydrated. To expose RNA targets, tissues underwent target retrieval in 1X Tris-EDTA (pH 9.0) at approximately 99°C in the steamer for 20 minutes, followed by 1 μg/ml Proteinase K digestion at 37°C for 15 minutes and post-fixation in 10% neutral buffered formalin. In situ hybridization was performed by applying the GeoMx RNA Probe Mix and incubating the slides overnight at 37°C in a humidified hybridization chamber. Stringent washes were performed twice to remove off-target probes. Finally, slides were blocked with Buffer W and incubated with morphology marker solution (SYTO13, PanCK, CD45) at room temperature for one hour to guide ROI selection after scanning on the GeoMx Digital Spatial Profiler. Oligonucleotide tags were cleaved from all selected ROIs and dispensed into individual wells in 96-well collection plates for subsequent library preparation.

Collected DSP aspirates from all ROIs were PCR-amplified using the GeoMx NGS Master Mix and GeoMx Seq Code Primer Mix to append unique dual i5/i7 indices and P5/P7 adapters for Illumina sequencing. Equal volumes of the PCR products from each well were pooled and then purified using Agencourt AMPure XP magnetic beads to remove residual primers. After dilution, the purified library was analyzed using the Tapestation for appropriate fragment size and quantified via qPCR to determine the optimal loading concentration for sequencing. All samples were finally sequenced using Illumina NovaSeq 6000 SP and NovaSeq X 10B in the FIMM Genomics NGS Sequencing unit at the University of Helsinki, supported by HiLIFE and Biocenter Finland, and preprocessed using the DRAGEN pipeline^38^.

##### 3.3 GeoMx data quality control and preprocessing

Raw DCC files originating from 1088 AOI collected as a part of this study were joined with 320 AOIs previously published in Launonen et al. 2024^15^ and processed together in all the subsequent steps.

DCC files were handled using the GeomxTools R package^39^, provided by Nanostring. Segment (AOI) and probe-level QC were performed as described in the package vignette. Overall, 48 AOIs were removed due to insufficient sequencing quality, and the remaining 1360 were further processed. Later, counts for the probes targeting the same gene were aggregated. Additional QC was performed, based on the limit of quantification (LOQ), as described in the GeomxTools vignette. For each segment, the Gene Detection Rate (GDR) parameter has been computed as the fraction of genes above LOQ in each segment. Segments were removed based on the GDR threshold of 0.03 (less than 3% of genes above LOQ). Since we expected a high level of variance in our dataset, the GDR threshold was set up to 0.05 (gene was removed if expression > LOQ in less than 5% of AOIs). 4503 genes and 180 AOIs were removed during this procedure, leaving a total number of 1180 AOIs and 14174 genes detected in the whole dataset with sufficient quality. The mean number of genes detected per AOI in the final dataset was equal to 5058.

Next, data were normalized using the Q3 quartile normalization method from the GeomxTools package, and dimensionality reduction was performed using t-distributed stochastic neighbor embedding (t-SNE) and UMAP, implemented with Rtsne^40^ and umap^41^ R packages.

Normalized data have been subjected to batch effect correction using the Harmony^42^ R package, after identifying the main batch number and secondary batch number (corresponding to mini-batches for samples handling during the wet lab procedure) as main sources of variance within the dataset (assessed with the PVCA method^43^).

##### 3.4 GeoMx and t-CycIF data pairing

Whole slide image alignment was achieved in QuPath^44^ through the Warpy plugin^45^, which performs affine registration, incorporating the moving image to the coordinate system of the base image. The resulting files were exported as pyramidal ome.tiff files to be analyzed in other platforms. For each ROI in the GeoMx slide, the corresponding coordinates on the t-CycIF slide were calculated, and all the cells identified in the t-CycIF slide (Material and Methods 2.2) were counted using the point.in.polygon() function from the sp R package^46^.

##### 3.5 GeoMx signal deconvolution

Since GeoMx data for each AOI contain a mixture of signals from each cell type within the selected tissue fragment, we deconvolved the signal to obtain cell-type-specific transcriptional profiles. To do that, we utilized 2 methods: 1) BayesPrism^47^ - designed for deconvolution of bulk RNAseq data, but validated on various spatial transcriptomics data, which, apart from the fraction of cell types, is able to compute the cell-type specific transcriptomics profiles; 2) SpatialDecon^48^ – a method designed specifically for GeoMx spatial transcriptomics data, returning fractions of given cell types.

Since both methods are based on reference scRNA data, we utilized the publicly available GSE266577 dataset, including 48 HGSC samples (n=22 pairs of IDS and PDS samples originating from the same patient, and n=6 unpaired IDS samples). This dataset was further down-sampled in a patient and cell-type-specific manner, generating the final dataset containing 31433 cells and 36571 genes. The downsampling procedure was performed by calculating the overall frequency_ratio parameter as mincell_nr/number of the least abundant cell type in the whole dataset, where min_cell_nr=100. Next, top_n=n*frequency_ratio cells per cell type and sample were selected, and tumor cells with less than 10 cells per patient were removed.

Due to the limitations of both algorithms in correctly predicting fractions of fine-grained immune cell types, and because less frequent cell types are not sufficiently represented in AOIs that typically contain 300 cells, we decided to cluster similar cell types into more general groups. Finally, as a reference for deconvolution algorithms, we used scRNA-seq profiles of: CD8+ T cells (Tem/Trm cytotoxic T cells), CD4+ T-cells (Tcm/Naive helper T cells), other T-cells (Regulatory T cells, Type 17 helper T cells), tumor cells, Macrophages/Monocytes (Classical monocytes, macrophages), Fibroblasts/Mesothelial cells, B cells (Memory B cells, Naive B cells, Plasma cells), Dendritic cells (DC1, DC2, Migratory DCs, pDC), NK cells (CD16-NK cells, CD16+ NK cells, NK cells), Mast cells, Endothelial cells and ‘other’ cells (ILC, Late erythroid cells). All initial cell type labels were obtained as described in Launonen et al., 2024^15^.

For BayesPrism method, we performed deconvolution according to their vignette, filtering out low complexity genes from the reference downsampled scRNA-seq dataset, using the above-mentioned cell groups as ‘cell types’, while detailed cell type labels were used as ‘cell states’ annotation (for tumor, cells originated from each patient were annotated as separate cell states). For SpatialDecon, we followed their vignette, using a custom profile matrix prepared using the downsampled scRNA-seq dataset. For both methods, we used raw scRNA-seq counts and raw and Q3 normalized GeoMx data for BayesPrism and SpatialDecon respectively.

Finally, BayesPrism deconvoluted expression profiles were normalized with Deseq2 VST method^49^ and batch corrected using Harmony^42^. The output of BayesPrism represents the mean expression values for all cells of a given type within a sample, so no further correction for different cell-type frequencies is needed to compare results.

Deconvolution results were validated by comparing the cell type fractions obtained from both methods and the cell types counted in the corresponding region of the adjacent t-CycIF slide (Materials and Methods 3.4, Supplementary Figure 2). We found sufficient correlation for the fractions of immune cell types (>0.8 between deconvolution methods, and >0.6 between deconvolution and t-CycIF), although in some cases the methods differed noticeably in the estimated number of tumor cells. Additionally, we validated cell type specific transcriptomics profiles obtained from BayesPrism, by demonstrating that the activity of the set of marker genes of the given cell type (assessed with single sample gene set enrichment analysis - ssGSEA) grows exponentially with the predicted cell type fraction for all abundant cell types (Tumor, Fibroblasts, Macrophages, CD8 T-cells, CD4 T-cells, DCs, Bcells), and is the highest in corresponding cell type specific deconvoluted transcriptomics profile (Supplementary Figure 3).

##### 3.6 GeoMx ROI clustering based on immune composition

To label ROIs based on their immune composition, we averaged the SpatialDecon AOI cell-type fractions per ROI. Next, we computed the relative abundance of each of main immune cell types (Macrophages/Monocytes, DCs, CD8+ T cells, CD4+Tcells, B-cells, and other immune cells comprising of other T cells and NK cells) to all immune cells in a given ROI, referred to as ‘immune fraction’, and used it to cluster the ROIs. The clustering has been performed using the GMM method from the mclust^50^ R package, and an optimal number of n=5 clusters was selected based on the elbow method (comparing the inertia and average silhouette score) and maximizing cluster separation in UMAP projection.

##### 3.7 GeoMx pathway activity analysis

We selected 267 signatures from the MSigDB database^51^ (Hallmark, GO:BP, KEGG:MEDICUS, PID, Reactome and Biocarta collections), as well as 14 signatures collected from the literature^52–55^ to compare their activities across AOIs. Selected pathways were related to the processes such as regulation of T cells and macrophage cell biology, and immune response; as well as key signaling pathways such as IFN, TNFα, TGFβ, IL2-JAK-STAT, IL-6, and IL-10. The full list of selected signatures can be found in Supplementary Table 4.

The activity of each gene signature has been calculated and scaled using ssGSEA method implemented in GSVA R package^56^.

3.8 GeoMx calculation of Differentially Expressed Genes (DEGs) and Gene Ontology (GO) enrichment

DEGs between AOIs of different cell type compositions were calculated based on deconvoluted cell-type specific transcriptomics profiles (see Materials and Methods 3.5). To diminish the noise, we excluded all AOIs with given cell type frequency (calculated with BayesPrism) lower than 0.01. DEGs were calculated using a linear mixed model (LMM) with SampleID as a cofounder (random slope in the model formula: ∼ ROI_type + (1 + ROI_type | SampleID)), as implemented in GeomxTools vignette (https://github.com/Nanostring-Biostats/GeomxTools).

GO overrepresentation analysis was performed with the enrichGO() function from the ClusterProfiler R package^57^ separately on identified over- and under-expressed genes (FDR <= 0.05 and log2FC > 0.5 or log2FC < -0.5). Identified GO terms were further clustered based on the semantic similarity of the terms with the reduceSimMatrix() function from the rrvgo R package^58^ (using 0.5 as similarity threshold) and filtered to remove all the parent terms. Remaining terms were ranked by Rich Factor and filtered to the one highest-ranked GO term per similarity cluster to represent only unique terms.

##### 3.9 GeoMx inference of Ligand-Receptor interactions

Ligand–receptor (LR) interaction analysis is implemented with the MultinicheNetR algorithm^59^, adapted for use with BayesPrism cell-type-specific deconvoluted profiles. Since both methods expect an scRNAseq dataset as an input, deconvoluted profiles must be joined into a pseudobulk scRNAseq expression matrix prior to analysis.

To prepare this input dataset, all the raw cell-type-specific deconvoluted profiles were joined together to form an expression matrix with each column representing the mean cell-type_AOI expression profile. That results in treating celltype_AOI as a ‘cell’ and AOI as a ‘sample’. Next, cell types whose frequency was lower than 0.01 in the given AOI were removed to reduce noise. This raw pseudobulk dataset was used as an input to MultiNicheNetR.

MultiNichenetR was run according to the package vignette (https://github.com/saeyslab/multinichenetr), using the following parameters: sample_id = AOI_id, min_cells = 1 (minimum number of AOIs containing given cell type – this parameter was low because cell types with low abundances were already removed), min_sample_prop = 0.25 and fraction_cutoff = 0.1 (for filtering genes treated as expressed in a given cell type). In addition, DGE genes between conditions were provided externally from previous DGE calculations (see Materials and Methods 3.7) to account for SampleID as a cofounding factor. Final output LR interaction list has been filtered using the following criteria: activity_scaled < -0.5 | activity_scaled > 0.5 for either up or down direction of regulation (to remove LR pairs with low activity of downstream genes), grouped by ligand and ranked by prioritization score.

##### 3.10 Construction and validation of Myelonet-specific transcriptional programs

Myelonet-specific transcriptional programs (lipid-il10-m2, tnfr1-tgfb, il2-vegf, il1-tnfr2-mhc2) have been constructed by collecting all genes from transcriptional signatures constituting Myelonet-specific transcriptional programs (Supplementary Table 4) and selecting those which found to be significantly overexpressed (FDR <= 0.01, log2FC >= 0.5) in Macrophage-dominated ROIs (lipid-il10-m2_dge, tnfr1-tgfb_dge, il2-vegf_dge, il1-tnfr2-mhc2_dge) (Supplementary Table 3). Next, we calculated ssGSEA activity scores for each of the constructed programs (as described in section 3.7) based on the Macrophage-specific deconvoluted transcriptomics profile per stroma AOI. AOI-level ssGSEA scores were matched to t-CycIF-detected cell-type nodes via overlaid cells to GeoMx ROIs in section 3.4, resulting in 652 matched node–AOI pairs (49 samples). Within each ROI, we aggregated the sizes of all Myeloid-labeled components (component_label == "Myeloids"; sum of component_size) and, separately, all other immune components. For each of the ssGSEA pathway scores, we then fitted an AOI-level linear mixed model with a random intercept per sample,

ssgsea_pw ∼ (sum Myeloid size)/100 + (sum other size)/100,

the other-immune term adjusting for overall immune load. Each pathway was z-standardized across the 652 stroma AOIs, so coefficients are reported as SD change per 100 aggregated Myeloid cells; 95% CIs were approximated as coefficient ± 1.96 SE. A linear rescale of the outcome leaves the Wald statistics unchanged, so the z-scored model reproduces the raw-model p-values and FDRs exactly. p-values were adjusted across the eight readouts using the Benjamini–Hochberg procedure. All models converged.

#### 4. Genomic Analyses

##### 4.1 Genomic homologous recombination deficiency scoring

HRD status of the tumor was assessed using the ovaHRDscar package (https://github.com/farkkilab/ovaHRDscar)^58,59,60^, based on loss of heterozygosity, telomeric allelic imbalances, and large-scale genome transitions derived from WGS data. Samples with ovaHRDscar >= 54 were defined as HRD-positive, and those with tumor purity <10% were excluded from the calculations.

### Statistical analysis

Statistical analyses were performed using R version 4 and python 3.13.3. The threshold for significance was set up as less or equal to 0.05 in all analyses. Statistical tests for the figures are indicated in the figure legends, and further statistical details of the analyses are provided in the corresponding method sections.

## Data Availability

All data produced in the present study are available upon reasonable request to the authors.

## Acknowledgements

This study was co-funded by the European Union (ERC, SPACE 101076096). Views and opinions expressed are, however, those of the author(s) only and do not necessarily reflect those of the European Union or the European Research Council. Neither the European Union nor the granting authority can be held responsible for them. In addition, this study was funded by the Sigrid Jusélius Foundation, the Research Council of Finland (grant numbers 1339805, 350396), the Cancer Society of Finland, and Ida Montinin Säätiö (A.S.). We also wish to thank the Helsinki Biobank, Institute for Molecular Medicine Finland FIMM Genomics Unit, Biomedicum Functional Genomics Unit, IMM Genomics NGS Sequencing unit at the University of Helsinki, supported by HiLIFE and Biocenter Finland, for the sequencing and primary analysis. The authors used generative AI to check the grammar of the manuscript, after which they reviewed the text.

## Declarations & statements

### Author contributions

**A.F.** conceptualized and supervised the study, interpreted results, and wrote the manuscript. **I.N.** and **A.S.** performed multimodal data integration, computational analyses, and wrote the manuscript. **I.N.** performed GeoMx transcriptomics analysis. **A.S.**, **E.R.**, and **A.J.** conducted t-CycIF image processing. **E.R.**, **Z.K.**, and **A.S.** performed spatial network analyses with SPACEstat. **E.R.** and **Z.L.** established image alignment and registration. **M.T.** performed patient-level statistical modeling and integration. **Z.L.** and **G.A.** performed GeoMx experimental workflows and ligand– receptor analysis, respectively. **S.S.** carried out t-CycIF experiments. **M.S.**, **U.-M.H.**, and **A.V.** provided clinical annotations, cohort curation, and biological sample preparation.

