## Supplementary material for "Myelonets define spatiotemporal immunosuppressive programs in ovarian cancer": Supplementary_Figures.pdf

### Supplementary Figure 1

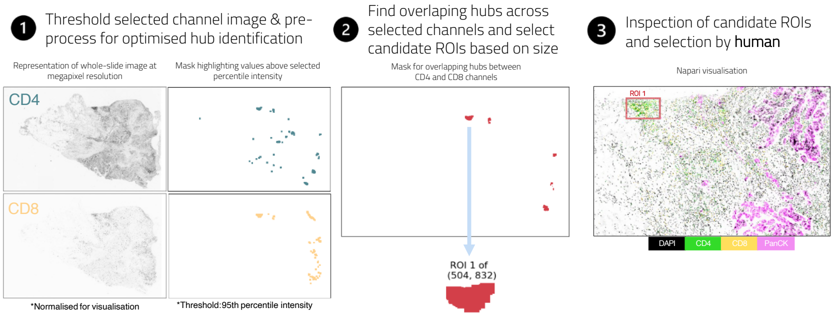


### Supplementary Figure 2

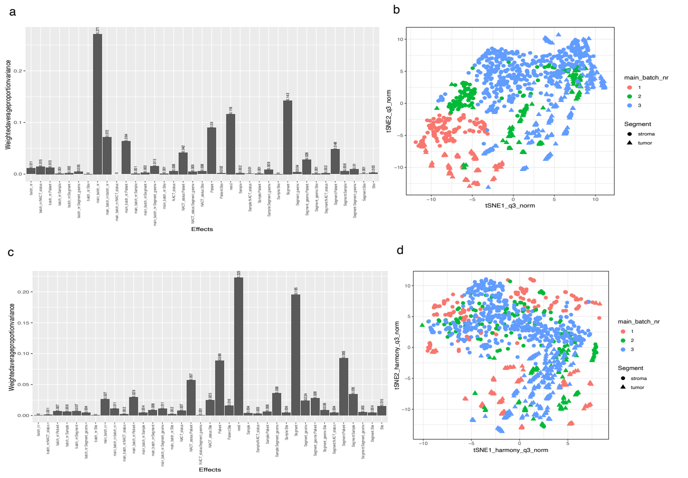


### Supplementary Figure 3

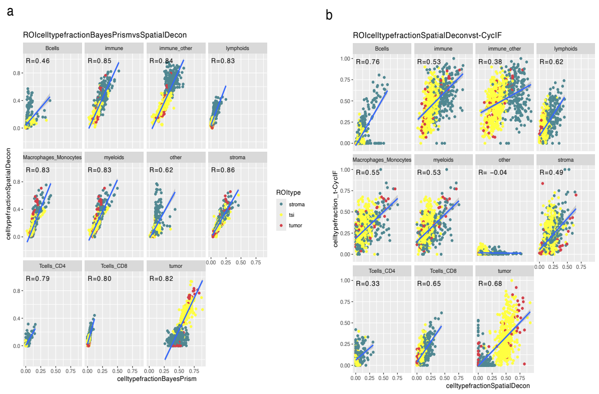


### Supplementary Figure 4

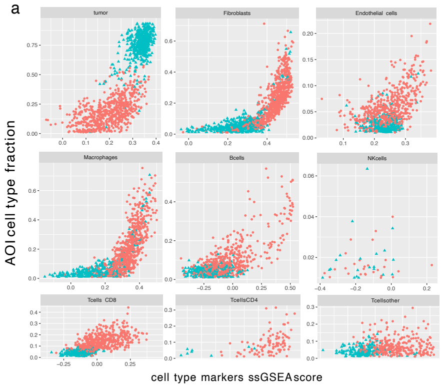


### Supplementary Figure 5

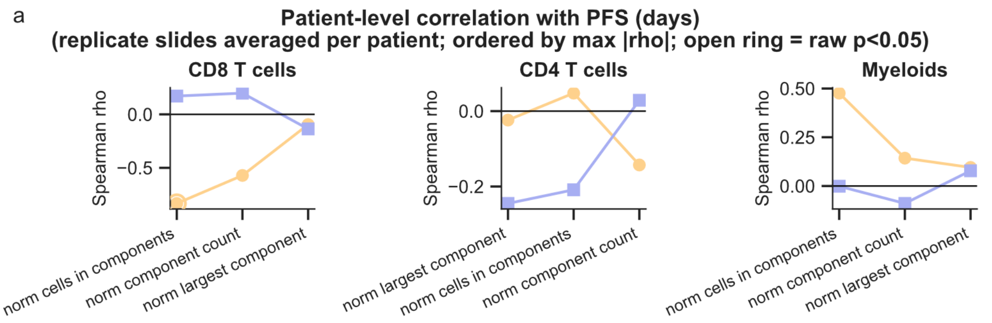


### Supplementary Figure 6

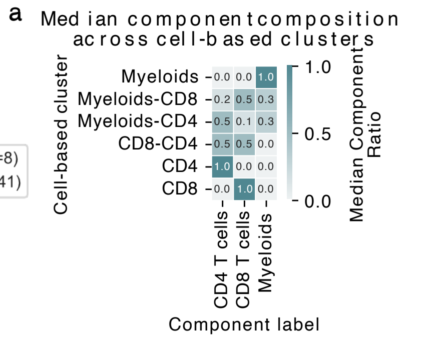


### Supplementary Figure 7

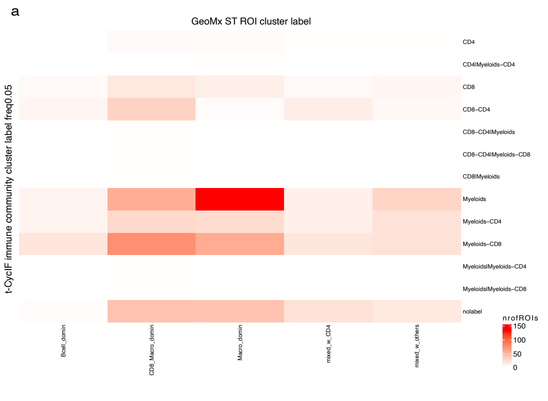


### Supplementary Figure 8

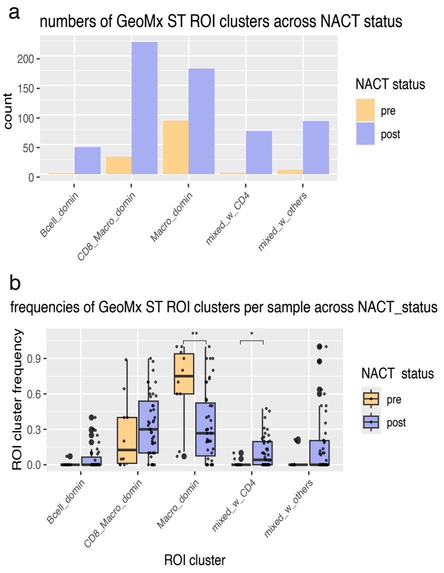


### Supplementary Figure 9

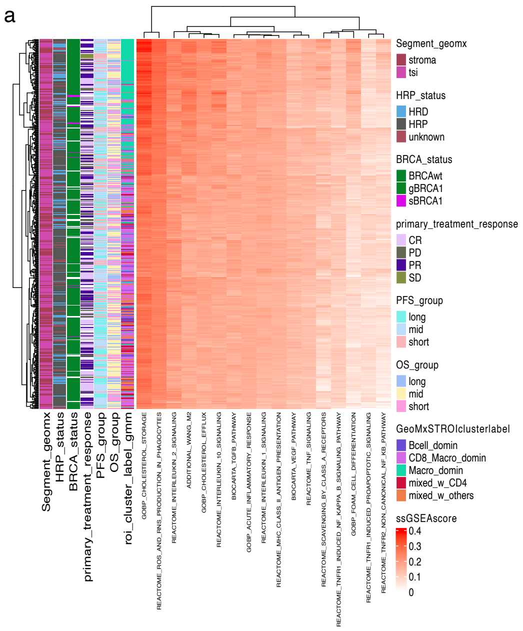
