## Supplementary material for "Myelonets define spatiotemporal immunosuppressive programs in ovarian cancer": Supplementary_materials_legends.pdf

SUPPLEMENTARY FIGURES LEGEND

SUPPLEMENTARY FIGURE 1

1A
Schematic overview of semi-automated t-CycIF guided ROI selection process. The tool has three key steps. (1) It preprocesses the selected channels to identify the immune aggregates. As examples, CD4 and CD8 channels are presented, where the final mask image shows potential immune aggregates of corresponding cell type. (2) The tool identifies the overlapping multi-cellular aggregates between selected channels and proposes candidate ROIs based on the size. (3) Finally, human is required to inspect candidate ROIs and select final tentative ROIs

SUPPLEMENTARY FIGURE 2

2A and 2C PVCA weighted average proportion variance of selected variables before (2A) and after (2B) harmony batch effect correction (n=1180 AOIs, n = 54 samples)

2C and 2D t-SNE projection for AOIs in GeoMx Spatial Transcriptomics data (n=1180 AOIs, n = 54 samples), coloured by main batch nr before (2C) and after (2D) harmony batch effect correction

SUPPLEMENTARY FIGURE 3

3A and 3B scatterplots with cell type frequencies per ROI (averaged across AOIs in given ROI) calculated with: BayesPrism and SpatialDecon deconvolution algorithms (3A) or SpatialDecon and cell type phenotyping on adjacent t-CycIF tissue slide (3B) with fitted linear model (y~x) (n=736 ROIs). R values represent Pearson correlation coefficient score, and color denotes different ROI types.

SUPPLEMENTARY FIGURE 4

4A scatterplot with AOI cell type fraction calculated with SpatialDecon deconvolution algorithm and ssGSEA score for signature of corresponding cell-type specific markers (Supplementary Table 4) calculated based on the full transcriptomics signal (n=1180 AOIs).

SUPPLEMENTARY FIGURE 5

5A Patient-level Spearman correlation between each component feature and PFS, in pre-treatment (chemo-naive, n=8 patients) and post-treatment (IDS, n=41 patients)

cohorts; replicate slides from the same patient and phase were averaged before testing. Each cell-type panel shows three component features – normalised cells in components, normalised counts, and largest-component size (normalised) – ordered by the maximum |rho| across the two cohorts. Chemo-naive shown as yellow circle-lines and IDS as purple square-lines; open rings mark raw p<0.05. No feature passed BH-FDR<0.05 within a screen; raw p<0.05 are highlighted as exploratory.

SUPPLEMENTARY FIGURE 6

6A Median fraction of each cell-based community cluster's resident immune cells belonging to CD4 T cells, CD8 T cells and Myeloids. For every community (n = 7,444), the fraction of its resident immune single cells of each type was computed (fractions within a community sum to 1); values shown are the median ratio per cluster, by cluster (rows) and cell type (columns). Pure clusters were strongly enriched for their defining cell type (Myeloids: 100% Myeloid; CD4: 100% CD4 T cell; CD8: 100% CD8 T cell), whereas mixed clusters showed graded, intermediate composition (Myeloids–CD8: 33% Myeloid/50% CD8; Myeloids–CD4: 31% Myeloid/50% CD4; CD8–CD4: 50% CD4/50% CD8). Cluster sizes: Myeloids n = 3,370; Myeloids–CD8 n = 1,149; CD8 n = 963; CD4 n = 735; Myeloids–CD4 n = 672; CD8–CD4 n = 555. Colour scale: median component ratio (0–1).

SUPPLEMENTARY FIGURE 7

7A Heatmap with comparison between ROI labels obtained with GeoMx ST ROI clustering based on immune composition calculated with SpatianDecon deconvolution algorithm and merged immune community cluster labels for all cells in corresponding region on adjacent t-CycIF tissue slide. t-CycIF community cluster labels were filtered to those representing at least 5% of all immune cells in each ROI. Since cells within certain regions in t-CycIF images were removed during QC procedure, only ROIs with at least 100 cells present in corresponding t-CycIF region, were compared (n= 678 ROIs)

SUPPLEMENTARY FIGURE 8

8A Numbers of ROI with given GeoMx ST ROI cluster label in pre and post-NACT samples

8B Frequencies of ROIs with given GeoMx ST ROI cluster label per sample. Asterisks represent significant p-values of paired wilcoxon rank-sum test between pre and post-NACT samples groups (* - p-value < 0.05, ** - p-value < 0.01)

SUPPLEMENTARY FIGURE 9

9A Clustered heatmap with ssGSEA scores for 18 selected signatures representing most important transcriptomic features of Myelonets communities (Supplementary Table 4) in all stromal AOIs of IDS samples (n = 398 AOIs, n=34 samples), calculated for Macrophage specific deconvoluted transcriptional profile. Annotations represent different clinical features of the samples.

### SUPPLEMENTARY TABLES DESCRIPTION

#### SUPPLEMENTARY TABLE 1

Information about cohort: tCycIF/GeoMx/WGS sample availability, sample chemotherapy treatment status (NACT_status), HRD status calculated with OvaHRDScar, Tumor Mutational burden (TMB) [mutation/Mb], Whole Genome duplication status (WGD), primary treatment response (CR – complete response, PR – partial response, PD – progressive disease, SD – stable disease), PFS group (short =< 350 days, long >= 602 days), OS group (short =< 613 days, long >= 1083 days). Main batch nr and t-CycIF Ab staining panel refers to antibodies used for sample staining (Supplementary Table 2)

#### SUPPLEMENTARY TABLE 2

Antibody panels used for t-cycIF and GeoMx sample staining, corresponding to t-CycIF AB panels, and GeoMx batches listed for each sample in Supplementary Table 1

#### SUPPLEMENTARY TABLE 3

Table containing list of DEGs computed based on Macrophage specific deconvoluted transcriptomics profile between Macrophage dominated ROI and all other ROI types, separately for tumor/stromal AOIs and pre/post NACT samples. Positive Estimate (log2FC) values indicated genes overexpressed in Macrophage dominated ROIs (Materials and Methods 3.8).

Table containing list of significantly overrepresented GO terms (p.adjust <= 0.05), based on list of significantly overexpressed genes in Macrophage dominated ROIs (FDR <= 0.01 and Estimate >= 0.5) in stromal AOIs of post NACT samples (Materials and Methods 3.8).

#### SUPPLEMENTARY TABLE 4

Immune signatures used for ssGSEA analysis

SUPPLEMENTARY TABLE 5

Table containing all combined results from ligand receptor inference with MultinicheNetR, calculated on Macrophage specific deconvoluted signal for Macrophage dominated vs all other ROI types for stromal AOI of IDS samples (Materials and Methods 3.9)

SUPPLEMENTARY TABLE 6

For each of myeloid-specific transcriptional programmes ssGSEA scores, a linear mixed model with a random intercept per sample was fitted on n_aois matched node–stroma AOI pairs (n_samples): z(ssgsea) ~ (sum of Myeloid component size)/100 + (sum of other-immune size)/100. Each outcome was z-standardized across the stroma AOIs. coef_sd_per100myeloid = model coefficient in SD units of the ssGSEA score per 100 aggregated Myeloid cells within the ROI, adjusted for other-immune aggregation; ci95_lo_sd/ci95_hi_sd = 95% CI (coefficient ± 1.96 SE); p_z = Wald p-value; fdr_q_z = Benjamini–Hochberg FDR across the eight readouts; sig_z = FDR-significant (q<0.05) flag; programme = myeloid-specific transcriptional programme (lipid metabolism-immunosuppression, inflammation-MHC-II, or other); iqr_myeloid_cells = interquartile range of summed Myeloid component size across the analyzed ROIs; coef_sd_per_IQR = coefficient rescaled per IQR of Myeloid aggregation (coef_sd_per100myeloid × iqr_myeloid_cells/100); median_stroma = pathway median ssgsea in the stroma AOIs; pct_of_median_per100 = raw coefficient per 100 cells expressed as % of the pathway median. Rows are ordered by descending coef_sd_per100myeloid.

SUPPLEMENTARY TABLE 7

Sheets define the hierarchical marker logic matrices used by Tribus to phenotype multiplexed imaging data, where values denote expected positive (1), negative/penalized (-1), or unconstrained/neutral (0) marker expression for a given cell type:

- Global_neg: First-tier classification partitioning cells into major compartments: Tumor (PanCK+), Stroma (Vimentin+/aSMA+ depending on available stroma marker in t-CycIF panel), and Immune (Iba1+, CD11c+, CD4+, or CD8a+).
- Stroma_neg & Tumor: Intermediate-tier logic resolving pure stromal or tumor populations from compartment-associated immune fractions.
- Immune, Stroma_Immune, & Tumor_Immune: Fine-grained immune lineage classification defining Iba1+, CD11c+ Myeloids and CD4+, CD8+ T cells across compartments.
